# Unraveling the spatial landscape of Dystrophinopathies: a transcriptomic approach to Becker and Duchenne muscular dystrophies

**DOI:** 10.1101/2025.05.30.25328395

**Authors:** Laura GM Heezen, Qirong Mao, Stefan Nicolau, Claudio Novella Rausell, Julia van der Weerd, Jan Kueckelhaus, Rasya Gokul Nath, Jordi Diaz-Manera, Hermien E Kan, Erik H Niks, Maaike van Putten, Annemieke Aartsma-Rus, Kevin M Flanigan, Ahmed Mahfouz, Pietro Spitali

## Abstract

Dystrophinopathies are caused by pathogenic variants in the *DMD* gene resulting in partial (Becker) or complete loss (Duchenne) of dystrophin. Becker (BMD) and Duchenne muscular dystrophy (DMD), are characterized by progressive muscle wasting, fatty replacement, fibrosis, and loss of function. To study histopathological changes, we used spatial transcriptomics to profile skeletal muscle biopsies of BMD, DMD patients and healthy controls (*N* = 4 per group). We estimated the proportion of cell types and their spatial localization across samples applying a deconvolution strategy using single-nuclei RNA-sequencing data. We identified genes enriched in fat patches and cell types such as fibroadipogenic progenitor cells (FAPs) in areas of active pathology. Using expression data of ligand receptor pairs, we highlight cell-cell communications leading to fibrotic and adipogenic lesions. Finally, analysis of gene expression gradients in areas of adjacent muscle and fat, allowed the identification of genes associated with muscle areas committed to become fat.

**Significance statement:** This study investigates the cellular and molecular changes that occur in muscles affected by Becker and Duchenne muscular dystrophy (BMD and DMD). These diseases are caused by mutations in the DMD gene, leading to muscle degeneration and the replacement of muscle tissue with fibrotic and fatty tissue causative for an early death. By using spatial transcriptomics, the researchers analyzed muscle biopsies from BMD, DMD patients, and healthy controls. They identified specific genes and cell types, such as fibroadipogenic progenitor cells, that are involved in disease progression. The study also revealed how different cells communicate with each other to drive muscle degeneration and fat accumulation. These findings provide new insights into the mechanisms of disease and potential targets for future therapies.

## Introduction

Becker muscular dystrophy (BMD) and Duchenne muscular dystrophy (DMD), collectively known as dystrophinopathies, are caused by mutations in the *DMD* gene, encoding the dystrophin protein. BMD patients experience a milder disease progression compared to DMD patients as a consequence of pathogenic variants leading to reduced levels of partially functional dystrophin, while dystrophin is severely reduced or absent in DMD (Gao & McNally, 2015).

Absence of dystrophin leads to loss of muscle fiber stability as the protein acts as a linker between the intracellular F-actin cytoskeleton and the extracellular matrix. As a consequence, muscle fibers are susceptible to contraction-induced damage, which in turn leads to chronic inflammation, cycles of impaired regeneration and degeneration, and finally, fibrofatty replacement of the muscle fibers (Duan et al., 2021).

Previous omics approaches, *in vitro* experiments, and histological studies uncovered gene expression signatures associated with muscle damage and the characteristic lesions typically observed in muscle biopsies of patients affected by dystrophinopathies. Such studies highlight the interplay of cell types required for successful regeneration in healthy skeletal muscle, and how the events leading to successful regeneration are not synchronized in dystrophic muscles.

These studies showed that, upon acute damage, a subpopulation of muscle resident stem cells, fibro-adipogenic progenitor cells (FAPs), proliferate and secrete trophic factors to stimulate myogenic activity of satellite cells. In a healthy situation, the damaged fibers are regenerated with help of these satellite cells, whereas the debris are removed by macrophages (Hao et al., 2022; Axelrod et al., 2021; Mázala et al., 2020) and the FAPs repair the connective tissue. The satellite cells proliferate and differentiate into myoblasts, then myocytes that fuse into multi-nucleated myotubes and eventually lead to the formation of new myofibers (Yin et al., 2013). However, the regenerative capacity of the myofibers is impaired in patients suffering from a dystrophinopathy when the satellite cells become exhausted due to chronic injury (Kodippili & Rudnicki, 2023; Duan et al., 2021). This, in turn, causes the FAPs to continue proliferating instead of ceasing their activity and differentiate into adipocytes or fibroblasts resulting in increased depositions of fat and fibrotic lesions in the tissue (Parker & Hamrick, 2021; Molina et al., 2021).

The mechanisms by which muscle tissue is replaced by fibrofatty tissue in dystrophinopathies, including the driving factors, underlying genes and cell types remain poorly understood. Understanding such processes is however essential to uncover the pathophysiology of the disease and to identify potential therapeutic targets as well as disease biomarkers. In this study, we used spatial transcriptomics to obtain paired histological and gene expression data with the aim of describing the changes in cell types and gene expression leading to histological lesions in patients affected by dystrophinopathies.

## Results

### A dystrophinopathy spatial transcriptomics dataset

In this study, we generated a spatial transcriptomics dataset from skeletal muscle biopsies to investigate dystrophinopathies. This included a total of *N* =12 samples (average age; standard deviation), these included *N* =4 BMD samples (9.75; 1.79), *N* =4 DMD samples (6.0; 1.87) and *N* =4 sex-matched controls (23.5; 5.5) (Table 1). Biopsies were obtained from lower and upper leg muscles in two sites (Nationwide Children’s Hospital, Columbus, Ohio, USA and Leiden University Medical Center, Leiden, The Netherlands) and the spatial datasets were generated using Visium from 10X Genomics.

**Table 1.**
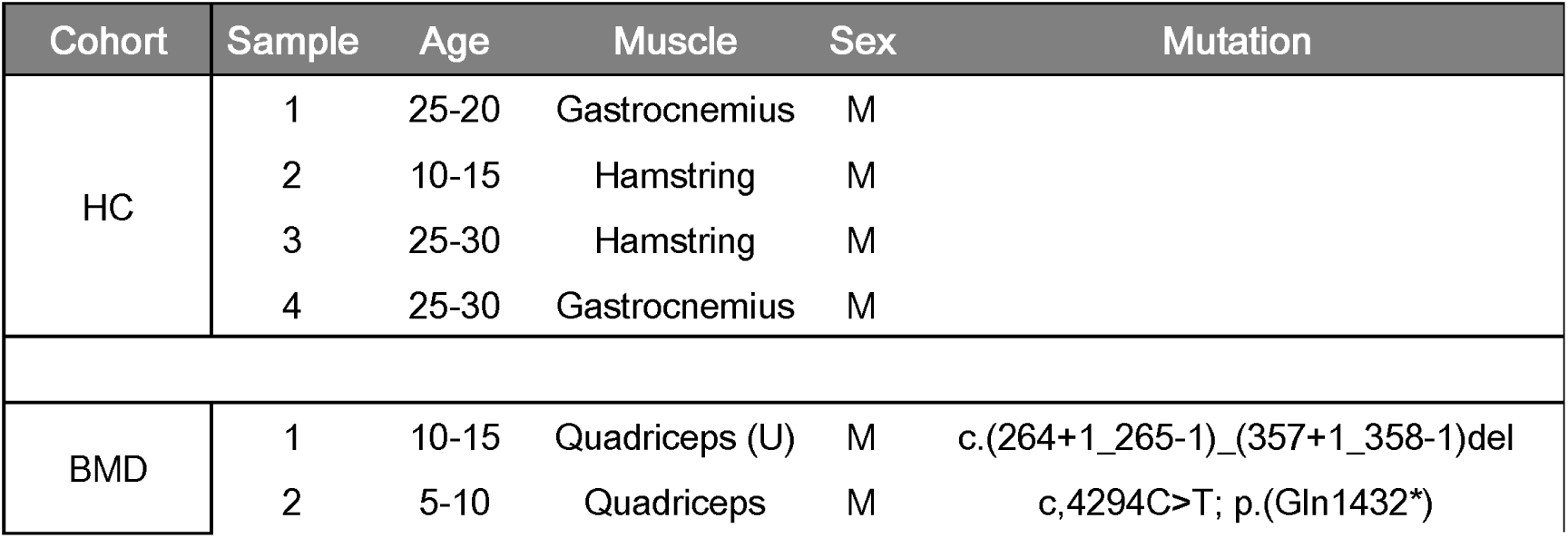
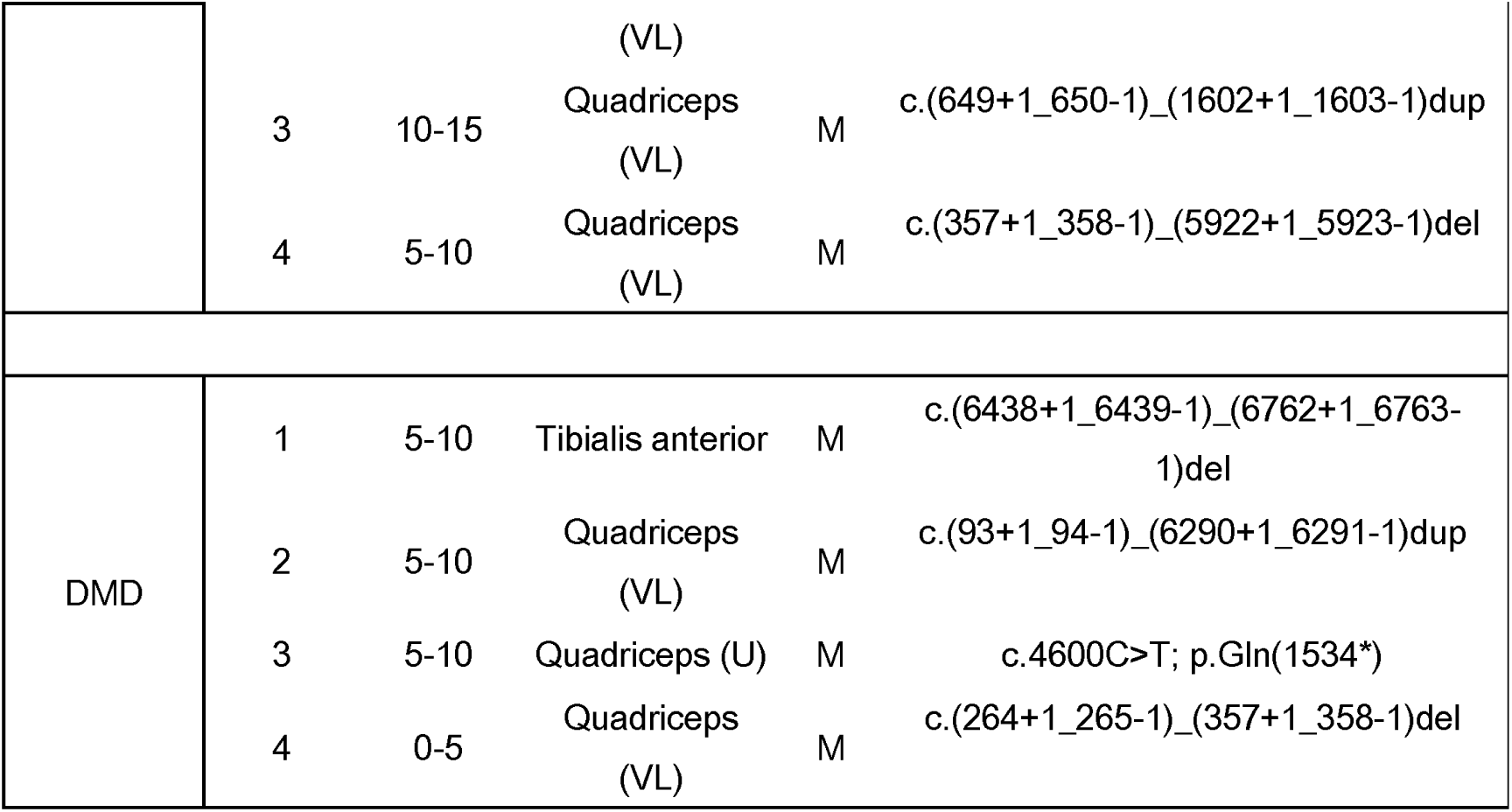
Patient characteristics of included samples in this study. (U = unknown which muscle head was biopsied; VL = vastus lateralis).

Using histological and transcriptomic data we annotated and compared sections across groups. To further elucidate the cell type contribution to histopathological lesions in the tissue, we utilized a deconvolution strategy using a single nuclei RNA-sequencing (snRNAseq) dataset (Suárez-Calvet et al., 2023) and assessed cell-cell interactions in these areas of active pathology. Lastly, we identified genes showing a spatial expression gradient surrounding intramuscular fat (Fig. 1a).

**Fig. 1.**
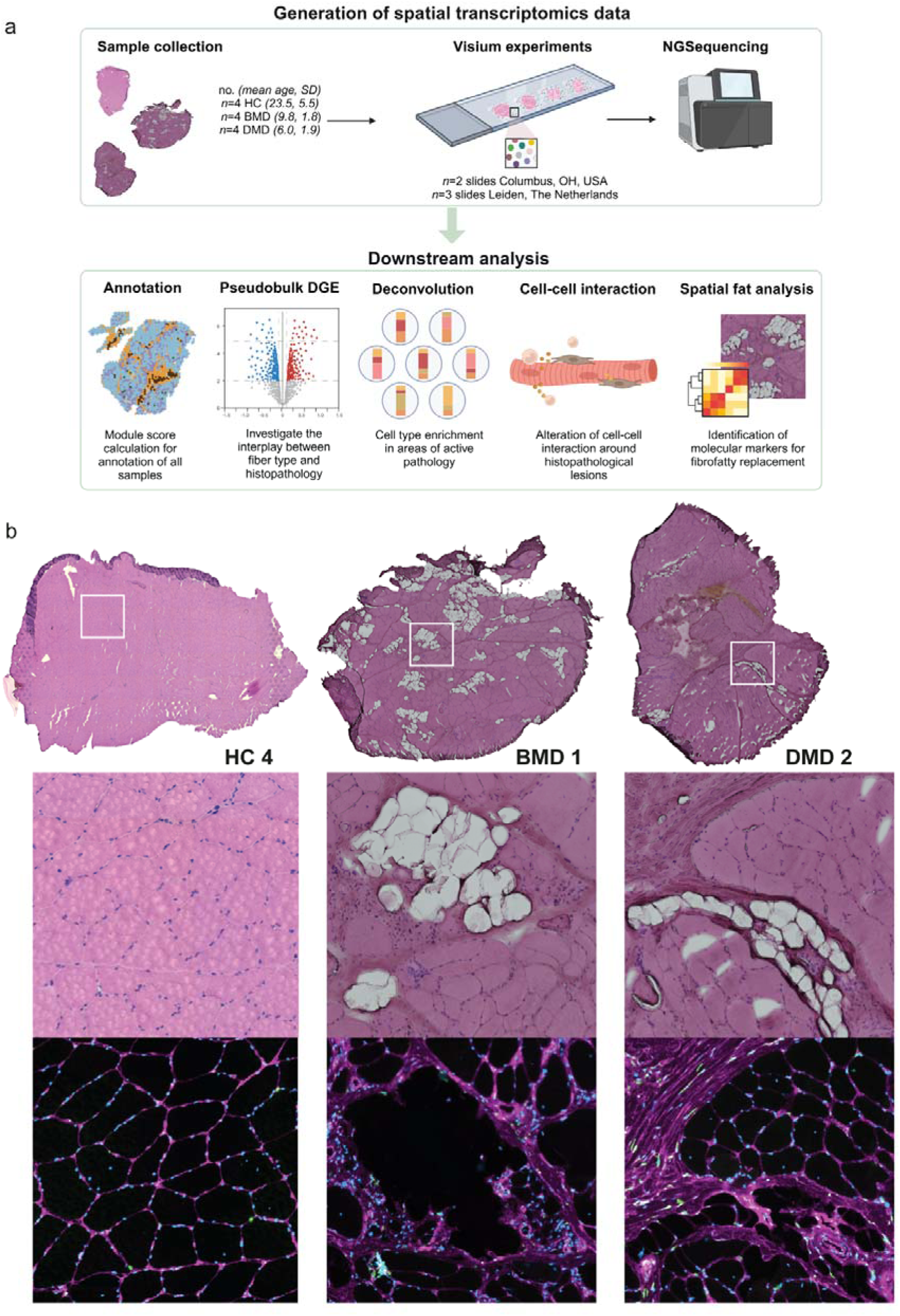
Study overview and representative histological features of included samples. (a) Schematic overview of the study and (b) representative HE images of one sample for each included group (HC, BMD and DMD) as well as a *FN1* staining for fibrosis.

Muscle biopsies of healthy controls were histologically more homogeneous in tissue composition, whereas dystrophinopathy samples showed a variety of histopathological lesions as reflected by the HE staining and confirmed by immunofluorescent staining of fibrosis and connective tissue marker FN1 on consecutive sections (Fig. 1b and Supplementary Figure 1-4). In both BMD and DMD, fibrosis, fat, inflammation, centralized nuclei and variation in fiber sizes were observed. The presence and extent of these histopathological lesions varied across samples (both within and across NMD).

Upon processing the samples, a total of 1,125,766,335 paired-end reads covering a total of 19,437 spots (8,183 spots HC; 5,305 spots BMD and 5,949 spots DMD) were obtained with a mean of 60,324 reads per spot (Table 2).

**Table 2.**
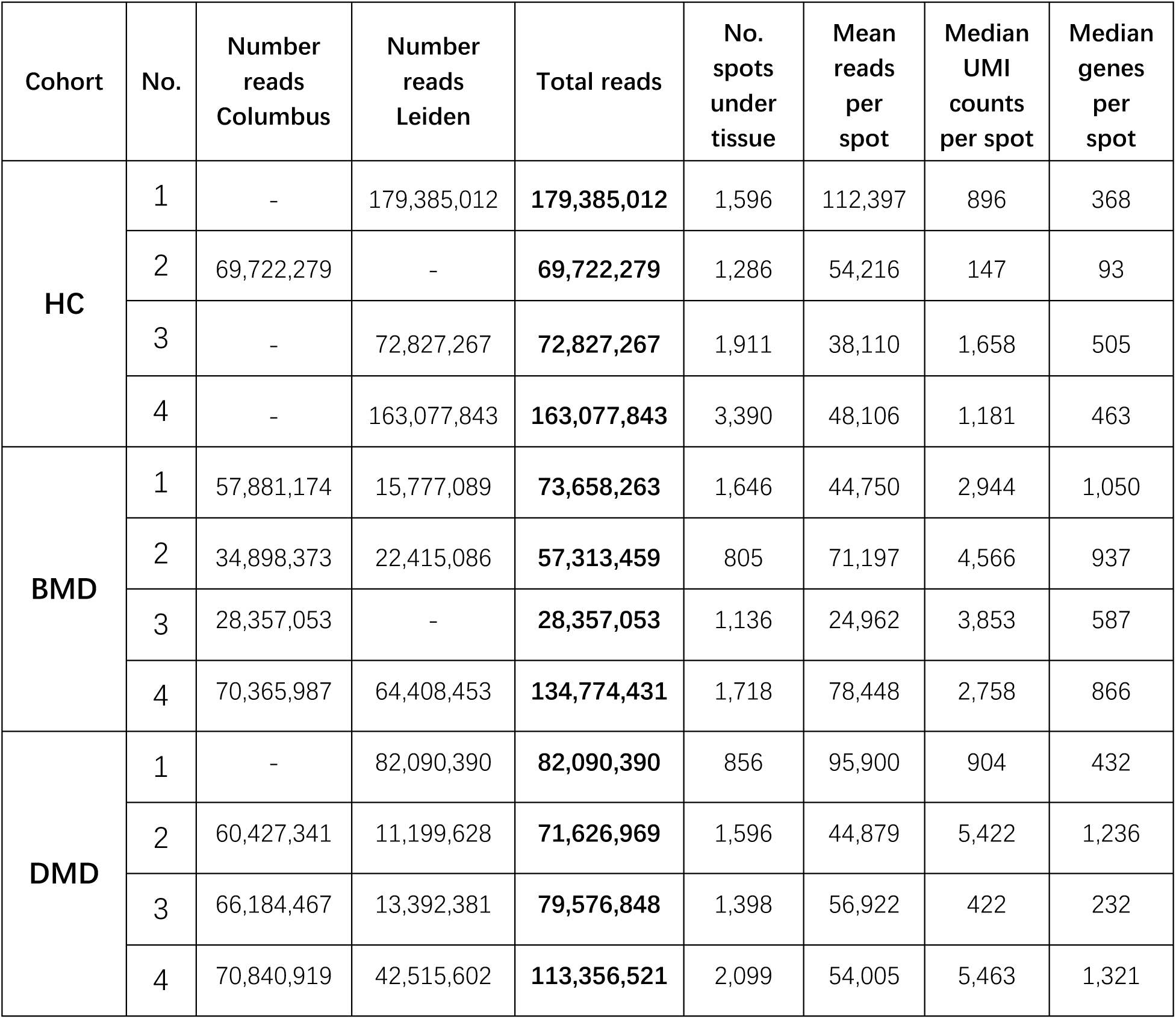
Overview of Next Generation Illumina sequencing results used in this study.

### Spatial transcriptomic data highlights a loss of 2X fibers in dystrophinopathies and increased connective tissue

To annotate Visium spots in each skeletal muscle sample, we employed a multi-step approach combining histological examination and expression of marker genes (Fig. 2a; Methods). First, fat spots were manually assigned based on assessment of the HE stained tissue images. Next, we used module scores to identify connective tissue spots was based on marker genes associated with the extracellular matrix, such as *COL1A1*, *COL1A2*, and *THBS4* (Supplementary Table 2). Similarly, we identified muscle fibers based on the module score and subsequently subdivided them into primary fiber types based on corresponding marker gene expression (Type I fibers: *MYH7*; Type IIa fibers: *MYH2*; Type IIx fibers: *MYH1*). Fig. 2d presents how the annotation of connective tissue and different fiber types aligns with the marker gene expression pattern in sample DMD 2.

**Fig. 2.**
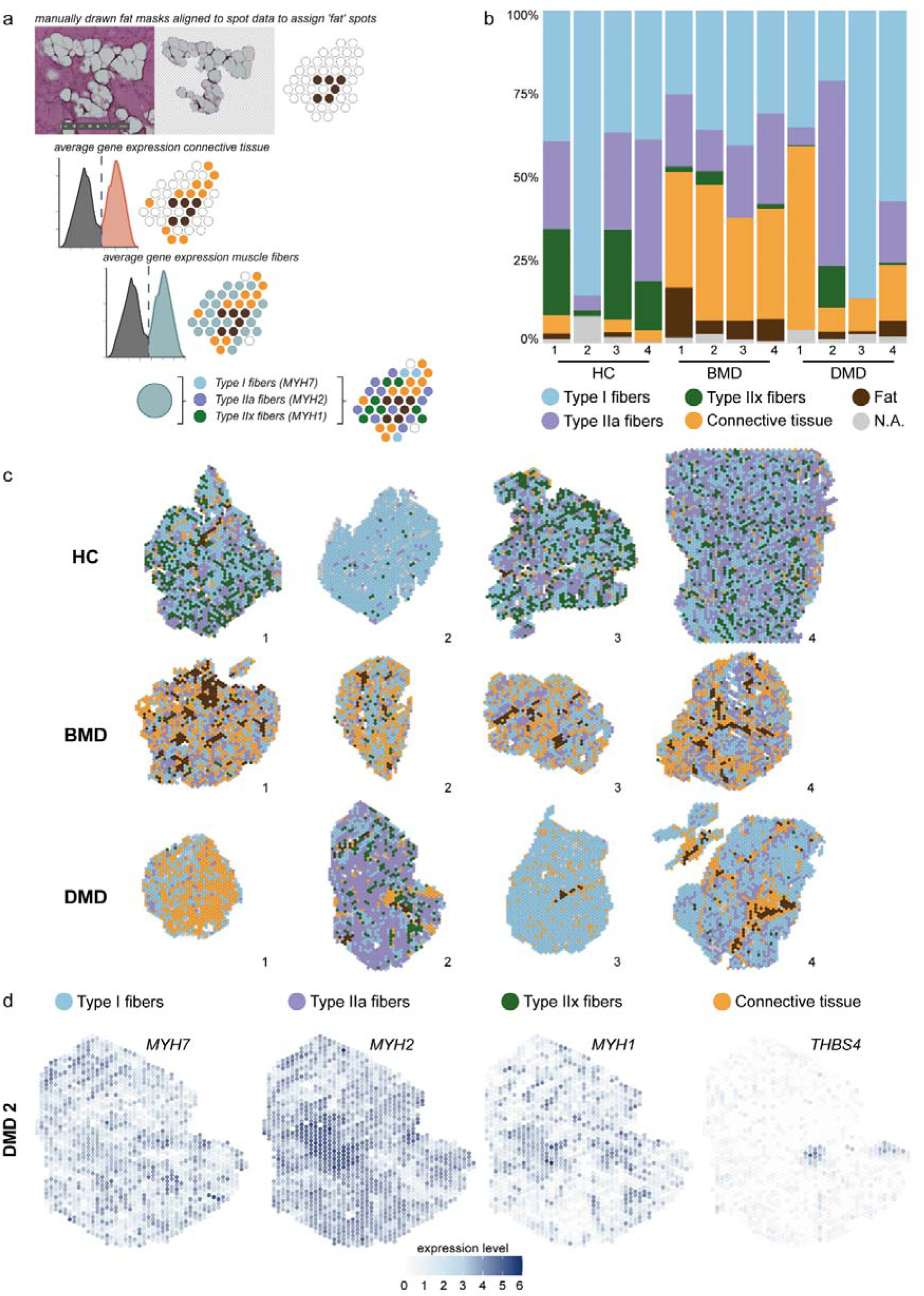
Annotation based on gene expression with module scored samples allows for comparison across groups and samples. (a) Assignment of modules to spots based on presence of fat masks, expression of connective tissue markers and expression of muscle markers. (b) Stacked barplot ranging from 0 to 100% representing the composition of models for all samples. (c) Spatial representation of module scored samples. (d) Example of gene expression of specific markers per module for DMD sample 2.

Annotation results showed that the tissue composition differs greatly across samples (Supplementary Table 3). As expected, myonuclei represent the predominant module in our samples (Fig. 2b-c; Table 3) with Type I fibers as the largest sub-module among the myonuclei, followed by Type 2A and Type 2X.

**Table 3.**
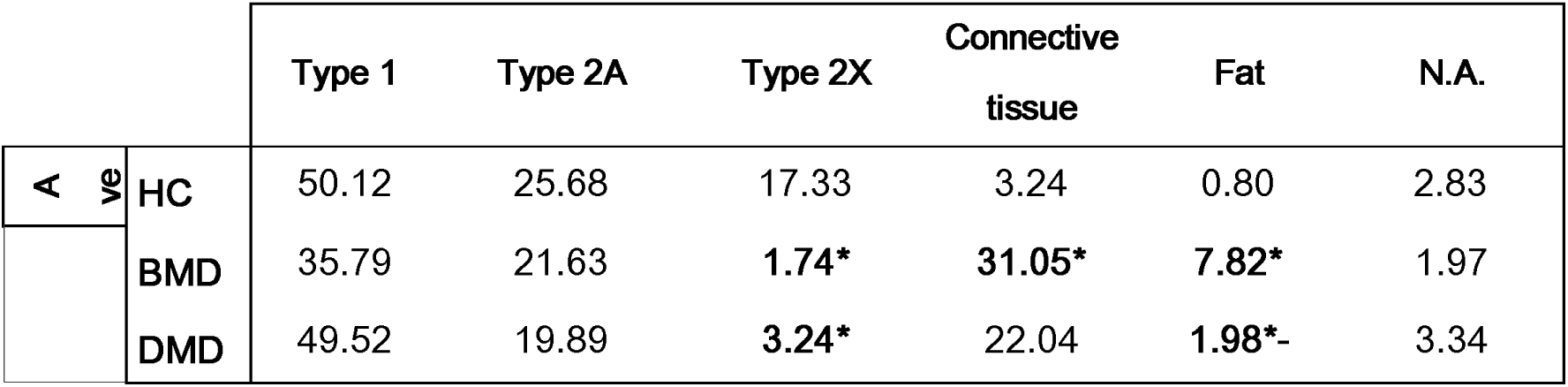
Average percentage of modules across HC, BMD and DMD samples. Significance of module annotation is calculated with a linear model comparing the three different groups with *N* =4 percentages per annotated module. Significance code: ‘*’ <0.05, with HC as a reference and ‘*-’ <0.05 with BMD as a reference. See Supplementary Table 4 for all linear model outcomes.

By comparing the different groups, we found that Type 2A had a higher average percentage in HC samples (25.68%) than in BMD and DMD samples (21.63% and 19.89%, respectively). Type 2X showed a significantly greater percentage in HC samples (17.33%) compared to BMD and DMD samples (1.74% and 3.24%, respectively). Additionally, Type 1 fibers in HC samples had a higher average percentage (50.42%) compared to BMD samples (35.79%), however not significant and there was no noticeable difference when compared to DMD samples (49.52%).

We also observed a higher percentage of connective tissue in the BMD and DMD samples (31.05% and 22.04%, respectively) compared to the HC samples (3.24%) with a significant difference for BMD samples only. A similar trend was evident in fat tissue, with increased proportions in BMD and DMD samples (7.82% and 1.98%, respectively) compared to HC samples (0.80%).

### Differential gene expression analysis linked to fiber type composition

To compare the transcriptomes of included samples, we performed pseudobulk pair-wise differential gene expression between groups (HC vs. BMD, HC vs. DMD and BMD vs. DMD) for each annotated region. Only a few differentially expressed genes were identified in the comparison between BMD and DMD samples, while the comparison between HC and dystrophic samples showed that multiple genes were differentially expressed in both BMD and DMD (Fig. 3a). Interestingly, comparison of the overall tissue with HC showed that BMD samples have larger differences compared to DMD, likely due to the higher occurrence of histological lesions observed in these particular sections in the BMD group. However, when looking into areas annotated as muscle fibers, a higher number of differentially expressed genes was observed in the DMD samples compared to BMD. This signature may reflect genes that are directly influenced by the absence of dystrophin or may relate to the more severe phenotype of muscle fibers in DMD (for all DGE genes across fibers, see Supplementary Table 5).

**Fig. 3.**
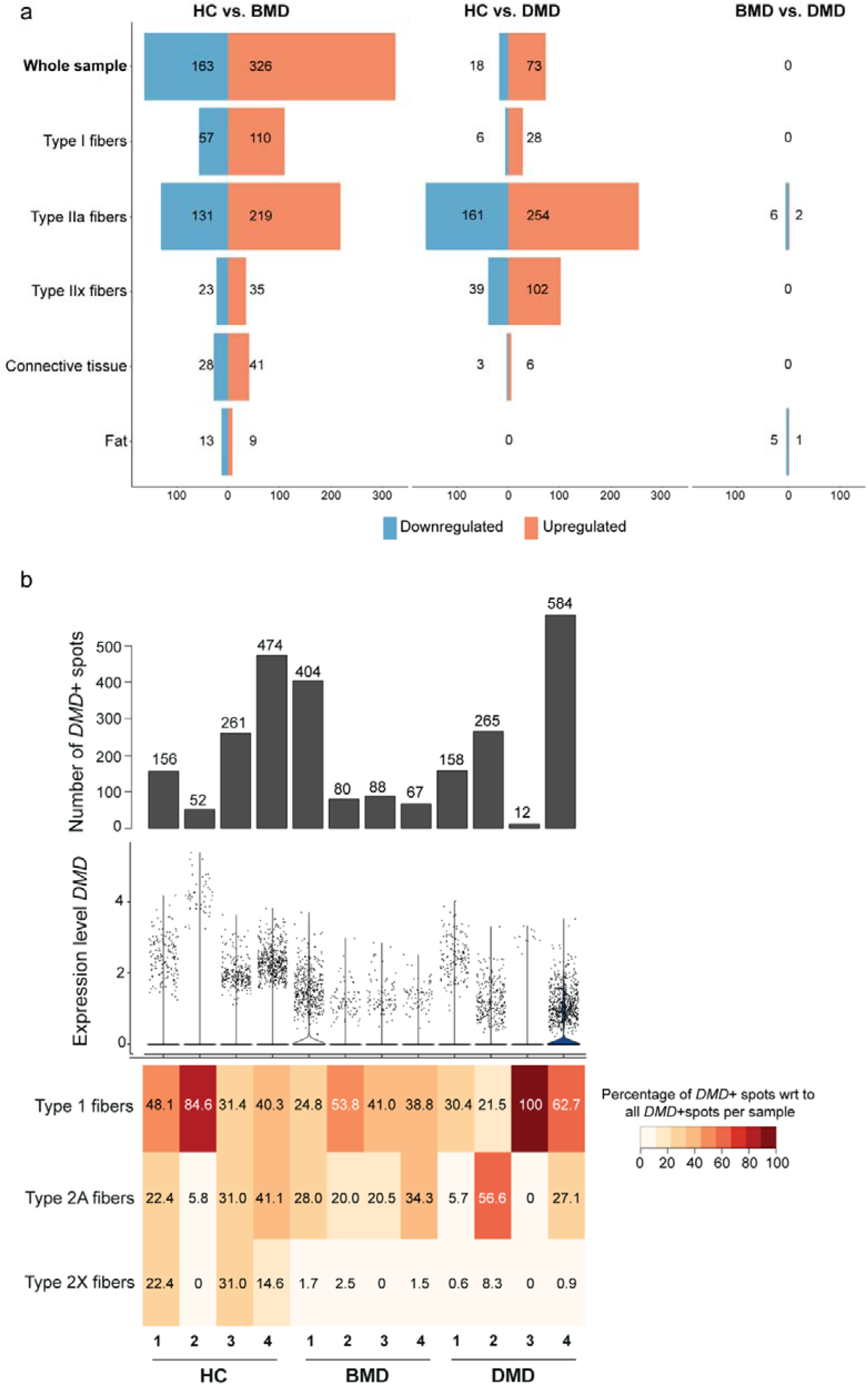
Differential gene expression (DGE) analysis highlights differences between HC and dystrophinopathies but not between dystrophinopathies. (a) Number of DGE genes (up- and downregulated) from pseudobulk analysis across samples and modules in pairwise comparisons between groups. (b) The absolute number of *DMD*+ spots in the samples, followed by the expression level of *DMD* in these samples and the percentage of *DMD*+ spots per assigned fiber type with regards to all *DMD*+ spots per sample.

When considering the ‘whole sample’ comparison, the top 5 upregulated genes in BMD vs. HC, ranked based on logFC were: *SPP1, HLA-DRB5, LINC01497, IGKC* and *CCL18*. These genes are mainly expressed in macrophages, endothelial cells and T-cells based on The Human Protein atlas (Karlsson et al., 2021). Whereas the top 5 significantly downregulated genes in BMD were: *ACTN3, AC024610.2, IRX3, MYH1* and *CFAP46*. These are mainly expressed in skeletal myocytes, fibroblasts, smooth muscle cells and a mix of cell types. These genes hint towards a more inflammatory profile in BMD skeletal muscle. By contrast, in DMD, the top 5 upregulated genes were: *MYH3, MYL4, MYH9, LINC01497* and *SHD*. According to The Human Protein atlas, these genes are mostly in skeletal myocytes, smooth muscle cells, endothelial cells, fibroblasts, macrophages (Karlsson et al., 2021). The top 5 downregulated genes were: *AC024610.2, CFAP46, NANOS1, MYOC* and *NR1D1*, which are mainly expressed in skeletal myocytes, smooth muscle cells and fibroblasts. While the BMD profile points mainly to an inflammatory signal, the DMD signature instead appears to be of a regenerative nature.

As discussed above, active pathology in dystrophinopathies is at the myofiber level where dystrophin is absent in DMD and reduced in BMD. *DMD* gene expression was variable in both amount of positive spots as well as the expression level (Fig. 3b). In general, dystrophin expression levels were higher in healthy individuals compared to DMD and BMD subjects (Fig. 3b). Considering the different fiber types, we observe that *DMD* expression tended to be higher in slow twitch (Type 1) fibers compared to fast twitch ones (Type 2A/2X), especially in the diseased samples (Fig. 3b, Supplementary Figure 5 for spatial plots). Higher dystrophin expression was found in DMD sample 1, which was however affected by the low number of muscle cells (especially type 2A and 2X) and the presence of an in-frame mutation in this patient despite the DMD phenotype (Fig. 3b).

### Enrichment of FAPs and adipocytes in histological lesions revealed by deconvolution

To determine the cell type contribution in each Visium spot, we used deconvolution based on a reference snRNAseq dataset, in which both healthy controls and DMD samples were sequenced (Fig. 4a; Suárez-Calvet et al., 2023). This reference dataset contained myonuclei originating from slow, fast and regenerating fibers as well as from smooth muscle cells. Moreover, non-myonuclei were also present such as adipocytes, FAPs, endothelial cells, satellite cells, and monocytic cells (Fig. 4a).

**Fig. 4.**
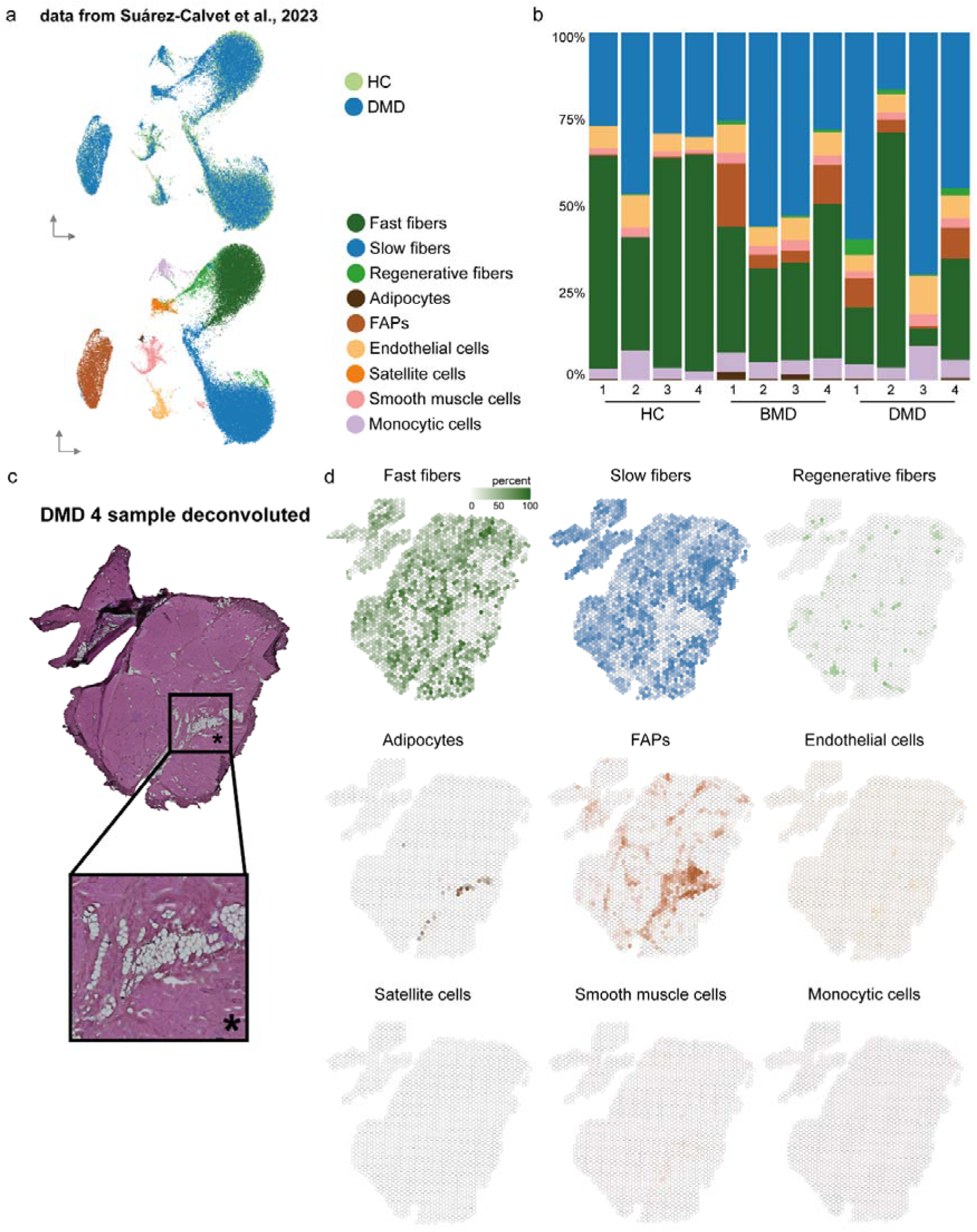
Deconvolution of samples shows cell type composition across groups and close to histopathological alterations. (a) Single nuclei RNA-seq reference dataset used for deconvolution with CARD derived from HC and DMD nuclei of different myo and non-myo nuclei. (b) Stacked barplot of cell type composition for all samples included in the dataset. (c) HE stained sample DMD 4 with zoomed-in area showing intramuscular fat and fibrosis. (d) Deconvoluted cell types per spot ranging from 0 to 100 percent.

The most prevalent cell types contributing to the transcriptomic profiles of the Visium spots were myonuclei, primarily slow and fast fibers, with large variation across samples (Fig. 4b). Dystrophic samples showed a lower percentage of fast fibers (33.93% and 29.54%) compared to HC samples (54.15%). Such changes were mirrored in the increased percentage of slow fibers in dystrophinopathy samples (average 40.42% and 47.59%) compared to the HC (average 33.12%) (Table 4). Changes in fiber type composition were evident in both deconvolution and module score analyses, suggesting an increased fragility of fast-twitch fibers in BMD/DMD compared to slow twitch fibers.

**Table 4.** Average percentage of deconvoluted cell types across HC, BMD and DMD samples. Significance of cell type difference calculated with a linear model comparing the three different groups with *N* =4 percentages per cell type. Significance code: ‘*’ <0.05. See Supplementary Table 7 for all linear model outcomes.

|  |  | Fast<br>fibers | Slow<br>fibers | Regenerative<br>fibers | Adipocytes | FAPs | Endothelial<br>cells | Satellite<br>cells | Smooth<br>muscle<br>cells | Monocytic<br>cells |
| --- | --- | --- | --- | --- | --- | --- | --- | --- | --- | --- |
| Average % | HC | 54.15 | 33.12 | 0.14 | 0.14 | 0.30 | 6.10 | 0.01 | 1.73 | 4.31 |
|  | BMD | 33.93 | 40.42 | 0.65 | **1.14 \*** | **9.16 \*** | 6.72 | 0.02 | **2.85 \*** | 5.10 |
|  | DMD | 29.54 | 47.59 | **2.09 \*** | 0.29 | 5.37 | 6.92 | 0.04 | 2.53 | 5.64 |

FAPs presence was higher in both BMD (average 9.16% which was a significant increase) and DMD (average 5.37%) samples compared to the HC (average 0.30%). Adipocytes were significantly increased in BMD (1.14%) compared to HC and DMD samples (0.14% and 0.29%), while regenerating fibers were significantly higher in DMD samples (2.09%) compared to BMD and HC samples (0.65% and 0.14%) (Fig. 4b; Table 4).

Average percentages were heterogeneous within each group, especially for BMD/DMD patients. As an example, some samples showed sizable increase in FAPs (BMD 1: 18.29% and DMD 4: 8.89%) while other biopsies characterized by less ongoing damage, showed lower FAPs estimates in the deconvolution results (e.g. FAPs in BMD 2: 3.84% and DMD 3: 0.72%). An overview of the percentages of all deconvoluted cell types by sample can be found in Supplementary Table 6.

Importantly, spatial data allows visualization of the deconvolution results on histological sections, enabling us to assess the presence of cell types at areas of histological lesions. For example, in DMD 4, adipocytes, identified through deconvolution, are mostly present in areas where fat patches are observed, and FAPs are present around fat patches and in areas of endomysial fibrosis (Fig. 4c-d). The zoomed image of the HE shows how the most severely affected region in the sample is characterized by substantial fibrofatty replacement (Fig. 4c-*), and how this region is enriched with FAPs and adipocytes, suggesting that FAPs are enriched in areas of active disease and adipocytes are present in areas of end stage pathology (Fig. 4d).

### Cell-cell communication around intramuscular fat depositions reveal crosstalk between FAPs, adipocytes and myofibers

To examine which cells are actively contributing to the histopathological lesions leading to the appearance of fat patches in dystrophic skeletal muscle, we performed cell-cell communication (CCC) analysis around areas of intramuscular fat depositions. This analysis allows for the detection of ligand receptor (L-R) pairs enriched in layers surrounding fatty infiltrations (Supplementary Table 8). Sample DMD 4 was used to visualize L-R pairs, while the other samples were used for further visualization (Supplementary Figure 8-9). Areas annotated as fat based on the HE images, were considered as layer 0 (L0); immediate neighboring spots were annotated as layer 1 (L1) and thereafter neighboring spots were annotated accordingly as layer 2 and 3 (L2, L3). All remaining spots were not assigned. Based on the previously described deconvolution results, every spot of the tissue was also assigned 1 cell type (the cell type with highest percentage presence per spot). Thereafter, we detected CCC across the different layers (layer 0 – layer 1; layer 1 – layer 2 and layer 2 – layer 3) based on expression of L-R pairs in at least 60 percent of the spots with at least two fold higher expression in these layers compared to the unassigned spots located far from fat patches (Fig. 5a; Supplementary Figure 8-9).

**Fig. 5.**
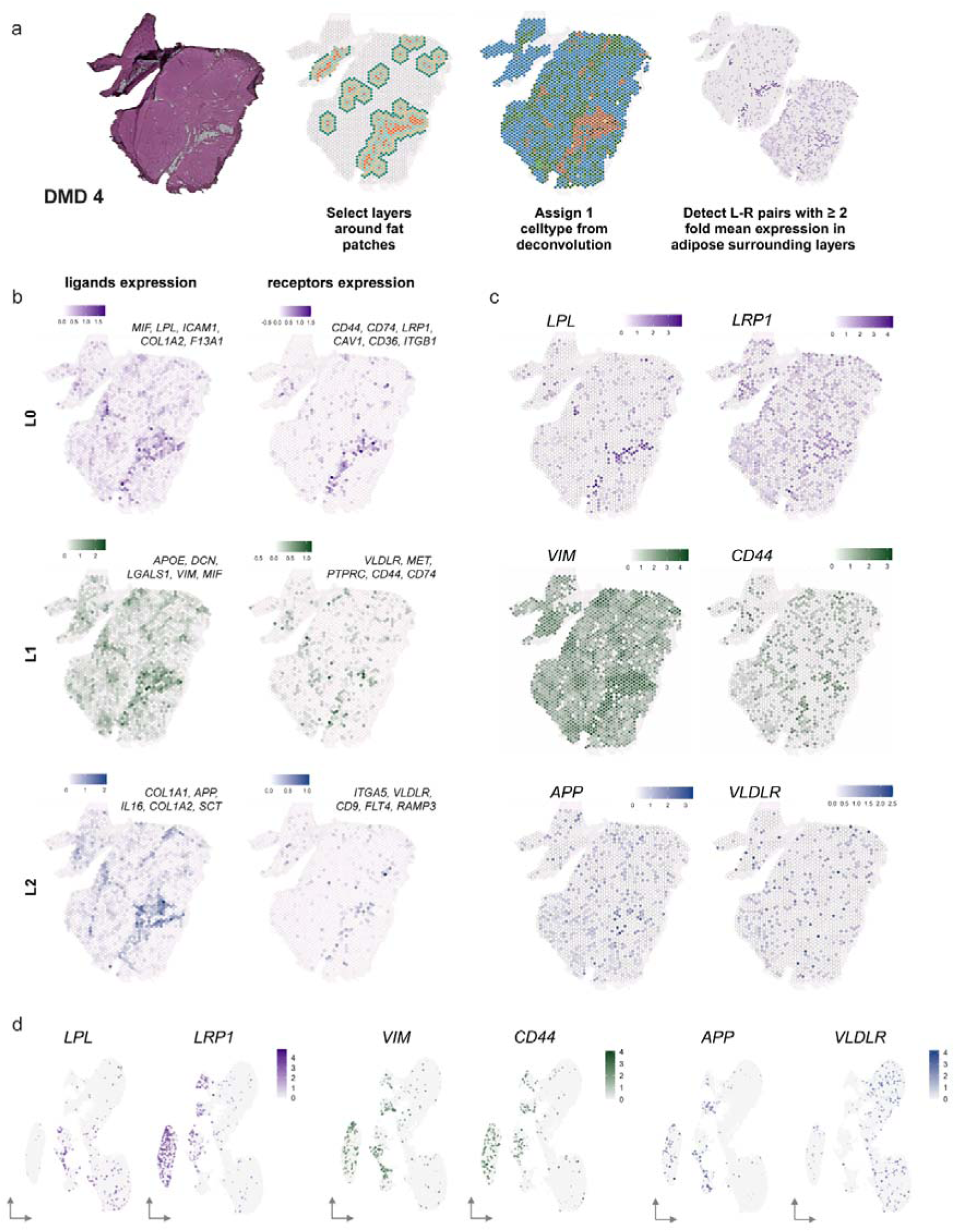
Cell-cell communication across layers surrounding intramuscular fat infiltration. (a) The set-up of layer selection, cell type assignment before detection of ligand-receptor (L-R) pairs across the layers. (b) Combined expression score of top 5 ligands and accompanied receptors across different layers (L0, L1 and L2). (c) Expression of one highlighted L-R pair per layer spatially plotted. (d) The expression of highlighted L-R pairs plotted in snRNAseq data pinpointing to potential sender and receiver cell types.

L-R pairs (L^R) enriched between L0 and L1 (meaning closest to the fat) were: *MIF^CD44/CD74, LPL^LRP1, ICAM1^CAV1, COL1A2^CD36* and *F13A1^ITGB1*. A combined, average expression score shows that the expression of these ligands and receptors was indeed localized in the areas surrounding the fat patches (Fig. 5b-L0). In the *LPL^LRP1* pair, *LPL* was specifically expressed in the adipocytes (senders) and *LRP1* was expressed in the neighboring layer enriched in FAPs (receivers) bordering the fat patch (Fig. 5c-L0). Confirmation of sender and receiver cell types was obtained from the snRNAseq data, where adipocytes were shown to express *LPL* and FAPs showed expression of *LRP1* (Fig. 5d). L-R pairs (L^R) that were enriched in the communication between L1 and L2 cells were: *APOE^VLDLR, DCN^MET, LGALS1^PTPRC, VIM^CD44, MIF^CD44/CD74*. Spatial plots of the average expression of these ligands and receptors highlight areas surrounding the fat patch, but also reflect areas where fibrotic tissue deposition was observed (based on HE images) and that are enriched in FAPs (deconvolution results; Fig. 4d) (Fig. 5b-L1). The senders in particular seemed to colocalize with areas enriched in FAPs. Spatial plots of the *VIM^CD44* L-R pair showed the that the pair is specific for areas surrounding histological lesions, as *VIM* expression was visible throughout the section, with the highest expression surrounding histological lesions, where the *CD44* receptor was present to capture the ligand (Fig. 5c-L1). Expression in the snRNAseq dataset suggests the senders to be the FAPs and endothelial cells, and receivers to be FAPs, satellite cells and smooth muscle cells (Fig. 5d). Lastly, L-R pairs (L^R) that were detected more distant from the fat between L2 to L3, were: *COL1A1^ITGA5, APP^VLDLR, IL16^CD9, COL1A2^FLT4, SCT^RAMP3*. Here, the average expression of the L-R pairs was outside of the fat patches and more in the fibrotic and surrounding areas (Fig. 5b-L2). The *APP^VLDLR* pair (Fig. 5c-L2), shows that *APP* was expressed by a combination of FAPs, satellite cells and endothelial cells, whereas *VLDLR* was expressed mostly by muscle fibers (slow and fast) (Fig. 5d). As FAPs were central in all L-R pairs, we re-clustered the snRNAseq data to identify whether specific FAPs subtypes were enriched in proximity to tissue lesions. FAPs subcluster 19 (Supplementary Figure 10a-b) was particularly enriched in these L-R pairs. These FAPs expressed marker genes such as *COL3A1*, *CFD*, *GSN*, *APOD*, *C3* and *CXCL14*, which have been previously connected to both adipogenic and fibrogenic commitment (Supplementary Figure 10c).

### Characterizing spatially-relevant adipose tissue markers

Substitution of muscle with adipose tissue is considered a hallmark of dystrophinopathies and the end histological stage of the pathological process. Importantly, muscle fat fraction is considered to be a prognostic biomarker for patients suffering from a dystrophinopathy. In order to identify marker genes of adipose tissue in dystrophinopathies, we assessed what genes were enriched in areas of muscle substituted by fat and what genes showed a gradient towards fat patches.

We selected dystrophinopathy samples (*N* = 3 BMD, and *N* = 2 DMD) with substantial fat infiltration, and selected spots that aligned with adipose tissue on the HE images. We then compared the gene expression signature in these areas with all ‘non-fat’ spots (Fig. 6a). Fourteen of the top 20 markers (based on Log2FC) were shared across the BMD and DMD samples (Fig 6b; Supplementary Figure 11 for spatial expression patterns). Known adipocytes markers such as *ADIPOQ* and a lipogenic enzyme *SCD* were identified, in keeping with previous reports (Li et al., 2020; Suárez-Calvet et al., 2024; Paran et al., 2015). Most detected genes are well described and linked to lipid metabolism and adipogenesis, e.g. *PLIN1, PLIN4, SCD, LPL* and *FABP4 but* have not been associated with dystrophinopathies before (Okumura, 2011; Carr and Ahima, 2016; Liu et al., 2019; Kiens, 2006; Wang et al., 2022).

**Fig. 6.**
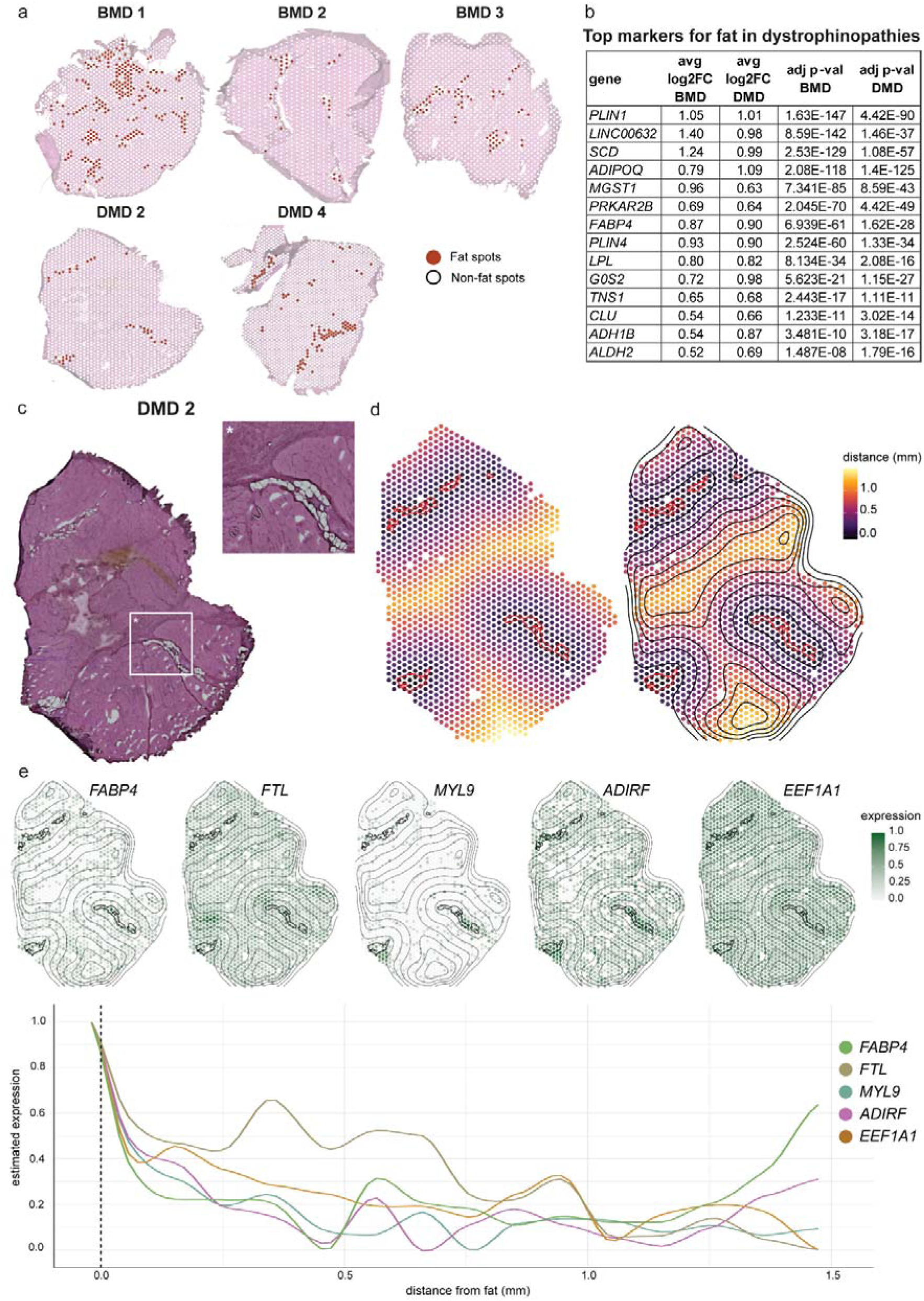
Identification of spatially relevant adipose tissue markers. (a) Selection of ‘fat’ spots (red) in BMD and DMD tissues for DGE analysis to all ‘non-fat’ spots (white) in these grouped samples. (b) Top (based on Log2FC) overlapping genes in fat spots in both BMD and DMD samples ranked by adjusted *P*-value. (c) HE stained image of DMD sample 2 with zoomed-in region surrounding fibrofatty infiltration. (d) Left: every spot is colored based on the distance from the fat masks (red line). Right: lines to visualize the gradient direction and orientation based on the distance from the fat masks. (e) Spatial and line plots of expression patterns of representative genes that have a decreasing expression pattern with increasing distance from the fat areas.

To identify genes with informative spatial patterning, especially surrounding borders of fatty infiltrations, we employed spatial fat analysis using SPATA2 (Kueckelhaus et al., 2024). This analysis takes the distance from the fat into account and aims to identify genes that may play an active role in the transition from muscle to fat, in areas where the muscle has not yet been replaced by adipose tissue. For this analysis, the same samples, i.e. those with substantial fat infiltration, were included (Fig. 6a). Fat masks were drawn on top of the HE image for every sample (Fig. 6c highlights DMD 2 as an example). Thereafter, the distances from the area of interest were calculated, and lines were added to visualize the distance based on the proximity to the fat masks (Fig. 6d). SPATA2 identified genes that align to pre-defined expression pattern such as linear descending, early descending, instantly descending, gradual peak, medium peak and small peak. Most identified genes fell in the descending instant, gradual or linear category (Supplementary Table 9). Genes that were identified in multiple samples were *FABP4, FTL, MYL9, ADIRF* and *EEF1A1* (Fig. 6e; Supplementary Figure 12), suggesting that these genes could be used to identify areas of muscle that are committed to transition towards fat but not yet visible in histology.

## Discussion

The dystrophinopathies Becker and Duchenne muscular dystrophy are characterized by tissue damage and clinical decline due to sub-optimal levels of dystrophin. In this study, we used spatial transcriptomics to identify gene expression signatures in relation to histological lesions in skeletal muscle biopsies of BMD and DMD patients. We mapped molecular changes to tissue alterations while retaining the spatial context, which is crucial for a better understanding of the histopathology and developing effective therapeutic interventions. The obtained data show how fast twitch fibers are particularly sensitive to reduced dystrophin levels and how FAPs are the drivers of tissue lesions leading to fibrotic and adipose tissue depositions. The spatial analysis allowed us to model patterns in tissue and interaction across cell types present in proximity, leading to a model of how such histological alterations come to exist.

Previous gene expression studies have enabled the identification of genes and pathways affected in dystrophinopathies using micro-arrays and bulk RNA-seq. Such analyses showed how lack of dystrophin (observed in DMD) leads to larger gene expression changes compared to samples with reduced levels of truncated dystrophin (as in BMD) (Xie et al., 2024). Our data show that histology and cell composition are the main determinants behind these gene expression signature. The gene expression signatures were similar in BMD and DMD when analogous histological areas and cell types were compared, with few differences in all tissue modules. However, comparing BMD and DMD samples to healthy controls showed that the gene expression signature is larger in fast twitch fibers (Type 2X and 2A) compared to slow twitch ones (Type 1). This comparison was however affected by the number of Type 2X fibers, which was severely reduced in both DMD and BMD, supporting earlier findings that fast twitch fibers are preferentially affected in dystrophinopathies (Webster et al., 1988). In line with this observation, the number of differentially expressed genes in Type 2X fibers was larger in DMD patients compared to BMD patients (both in comparison with healthy subjects), suggesting that higher levels of dystrophin can partially normalize the gene expression signature in fast twitch fibers. Interestingly, among the genes found to be differentially expressed only in DMD, we found genes involved in muscle regeneration (e.g *MYH3*, *MYH8* and *MYL4*), muscle contraction (*MYOM3*) supporting increased muscle damage in Type 2X fibers in DMD, the known genetic modifier (*LTBP4*) (Flanigan et al., 2013; Flanigan et al., 2023), and genes involved in lipid metabolism (*APOC1*, *APOE*).

A unifying pattern was found in both BMD and DMD, establishing how reduced levels or complete lack of dystrophin lead to tissue lesions. In both conditions, deposition of fibrotic and adipose tissue are observed. Deconvolution analysis showed that FAPs were enriched in areas of fibrosis and surrounding adipocytes, suggesting that active/ongoing pathology converges on the presence of FAPs, while the end-stage pathology is represented by the presence of adipocytes. While FAPs are key to successful muscle regeneration in healthy conditions, we observed these cells as the main drivers of histological lesions in both disorders. A recent omic study supports this finding FAP involvement with different expression patterns, but lacks spatial context (Ren et al., 2024). To understand how fibrotic and adipose tissues are deposited in muscle, we studied the co-expression of ligand-receptor pairs and the patterning of gene expression in contained areas. The goal of this analysis was to determine similarities and differences between ligand-receptor pairs leading to fibrotic and adipose lesions by relying on their proximity to fat patches. Cell-cell communication and expression in proximity to fat would provide information on the commitment of FAPs towards adipogenic lineage, while the same analyses further away from fat patches would allow to study FAPs committed towards fibrogenic lineage. The ligands and receptors we identified have previously been linked to lipid metabolism and homeostasis. As such, it is known that *LRP1* plays a role in lipid homeostasis as it binds lipoprotein lipase (*LPL*) (Verges et al., 2004). Moreover, in fibroblasts and smooth muscle cells, *LRP1* controls the trafficking of platelet-derived growth factor receptor β (*PDGFRβ*) which in turn is a marker linked to FAPs and pre-adipocytes (Wang et al., 2024). Disrupted expression of *LRP1* on smooth muscle cells has been shown to result in elevated levels of *PDGFRβ* expression, and atherosclerotic lesions and fibrosis (Boucher et al., 2003; Betsholtz et al., 2001). Further away from the fat patches, we identified interactions between *VIM* and *CD44*, which are known to mediate a highly conserved crosstalk across immune cells such as dendritic cells (DCs) and T-cells. It is thought that these interactions promote the antigen presentation and activation of autoreactive T-cells (Li et al., 2022; Bourguignon et al., 2004). Finally, on the furthest layer from the fat, we identify the *VLDLR* receptor. This receptor is present in the brain, adipose tissue, vascular endothelial cells, as well as heart and skeletal muscle (Reddy et al., 2011; Huang & Lee, 2022). The interaction between *APP* and *VLDLR* has mostly been studied in Alzheimer’s disease, a neurodegenerative disorder, and seems to be essential for normal brain functioning, neuronal development, as well as lipid metabolism (Oka et al., 1994; Reddy et al., 2011). Its role in neuromuscular disorders remains to be uncovered.

This study has a few limitations. Given that fewer muscle biopsies are now being performed for diagnostic purposes, we could only retrospectively access samples that were collected at different clinical sites and from different muscle groups. Moreover, the obtained healthy controls from The Hague were not completely healthy due to ACL surgery and were significantly older. Despite this limitation, the identification of common genes across muscles further supports the strength of the associations identified in this work. It is known that BMD is a heterogeneous disorder, patients included in this study showed symptoms at an early age and may be at the severe end of the spectrum, making them more DMD-like. The heterogeneity of the biopsies in the DMD group was larger than in BMD group, with two biopsies showing less prominent tissue lesions compared to the other six dystrophinopathy samples: one biopsy was rather small and showed no adipose tissue deposition and a second biopsy was enriched in slow twitch fibers, which are less prone to damage, showed less pronounced histopathology. Despite the limitations in this group, we could identify a severity signature, especially in fast twitch fibers, supporting the incremental fragility of fibers completely lacking dystrophin (as in DMD) compared to fibers presenting reduced dystrophin levels (as in BMD). Importantly, this comparison allowed us to propose genes that relate to dystrophin dosage/expression and could be used to monitor the effects of dystrophin reintroduction by dystrophin restoring therapies. Finally, the resolution of the Visium data did not allow us to study the spatial expression of single cells and we used a deconvolution strategy to estimate cell types at spatial locations. Future work should aim to bridge this gap and use spatial transcriptomic technologies that allow to study individual cells in histological sections. Nevertheless, the transcriptome-wide coverage allowed for the identification of novel markers (e.g. *LPL, LRP1, VLDLR, APP, PLIN1, FTL,* etc.) which would normally not be included in targeted panels due to their previously unknown link to dystrophinopathies.

To conclude, this is the first human spatial transcriptomic study on a genetically inherited muscle disease. The analysis enabled the identification of marker genes of histological lesions such as adipose tissue deposition and to propose genes and cell types involved in the transition of muscle to fat. Higher resolution studies using high resolution spatial technologies will enable to clarify the net effect of cell types in active pathology niche.

## Methods

### Ethical statement

The LUMC Toetsing Commissie for Neurologische ziekten evaluated the research protocol (B20.001) and approved with number 3.4258/010/FB/jr. Informed consent forms were obtained from one DMD patient and his legal representatives. The healthy controls included in this study from the Netherlands were obtained in The Hague, obtained from individuals undergoing anterior cruciate ligament (ACL) surgery. For these subjects (*N* = 3), the study was approved by the local Medical Ethical Review Board of The Hague Zuid-West and the Erasmus Medical Centre and conducted in accordance with the ethical standards stated in the 1964 Declaration of Helsinki and its later amendments (ABR number: NL54081.098.16). All other subjects (*N* = 1 healthy control, *N* = 3 DMD and *N* = 4 BMD) were included in the United States of America and this study was approved by the Nationwide Children’s Hospital Institutional Review Board (0502HSE046 and IRB14-00719). All subjects or their guardians provided written informed consent prior to participation. Patients were included in their respective dystrophinopathy group based on clinical diagnosis.

Skeletal muscle biopsies were obtained for healthy controls in the Netherlands after ACL construction. These biopsies were taken by percutaneous biopsy (modified Bergstrom, 1975) using a minimally invasive biopsy needle from gastrocnemius lateralis (GL) rectus femoris (RF), vastus lateralis (VL), and vastus medialis (VM). All biopsies were immediately frozen in liquid nitrogen and were kept at –80 °C until further processing. For all biopsies included from the United States of America, patient muscle samples were collected from the quadriceps muscle, while the healthy control sample was collected from the hamstrings. All samples were frozen in isopentane chilled with liquid nitrogen, according to standard techniques.

For the present study, we selected samples containing a combination of muscle fibers, adipose tissue, and fibrous connective tissue.

### Spatial Visium experiments

All biopsies (*N* = 12) were fresh frozen upon collection and cryosectioned in cross-sections of 10 µm thickness at −22 °C using a CryoStar NX70 cryostat (Thermo Scientific, Waltham, MA, USA) before being placed on one of the 5 Visium Spatial Gene Expression slides (PN: 2000233, 10X Genomics) in Leiden or Columbus (For detailed overview see Supplementary Table 1). The cross-sections were adhered to the slide by warming the back of the slides and were stored at −80 °C until further processing. The Visium Spatial Gene Expression slides were processed according to the manufacturer’s protocols. The slides were transferred on dry-ice to the PCR machine with slide adaptor and warmed to 37 °C for 1 minute prior to fixation in ice-cold methanol for 30 minutes at −20 °C. Slides were then incubated with isopropanol at room temperature (RT) for 1 minute after which the hematoxylin and eosin (HE) staining (Agilent Technologies, Santa Clara, CA, USA; Sigma-Aldrich, Burlington, MA, USA) was performed following 10X’ protocol. Images of the capture areas were taken using the ZEISS Axio Scan.Z1 slide Scanner (Carl Zeiss LTD, CAM, UK) with a 20x objective in Leiden and the Nikon Eclipse Ti2-E (Nikon Instruments, Melville, USA) with a Plan Apo λ 20x objective and a Nikon DS-Ri2 color camera (final resolution 0.37µm/pixel) in Columbus. Images were exported as TIFF files for further processing and alignment to the sequencing data. After imaging, permeabilization of the tissue was performed for 15 minutes. The released poly-adenylated mRNA from the tissue sections was then captured by the barcoded primers on the Visium slide. Through reverse transcription, in the presence of template-switching oligonucleotides, the captured mRNA was converted to spatially barcoded, full-length cDNA. Thereafter, second strand synthesis was performed. Finally, cDNA was released from the capture areas on the slides by denaturation and subsequently used for PCR amplification. The total number of cycles ranged between 16 and 23 and was determined based on Cq values (see Supplementary Table 1). Afterwards, enzymatic fragmentation and size selection (using SPRI beads) was performed for optimization of the cDNA amplicon size. Finally, through end repair, A-tailing, adaptor ligation and PCR amplification, P5, P7, i7 and i5 sample indexes, and TruSeq Read 2 adapter sequences were added to generate a sequencing ready indexed library. The spatial gene expression libraries were sequenced on an Illumina NovaSeq6000 with a target of ∽125 million Paired-End reads per sample, or 50,000 reads per spot for each sample.

### Drawing masks on top of HE stained images

Masks were manually made to annotate fat, gaps, and folds in the tissue. Masks were then aligned to the sequencing data. This was done in Photoshop with the image that was used in SpaceRanger. After duplicating the layer, the wand tool or magic wand tool were used to select the region of interest on the Visium image. For each area annotated as fat, a separate mask was made. Each region of interest was selected with the ‘select and mask’ function, with transparency set at 100% and saved as a new PNG document. Masks were then loaded into the Seurat object.

### Immunofluorescent FN1 staining

To visualize fibrotic tissue in skeletal muscle biopsies, serial sections were collected immediately after the Visium section (5 μm thick) on glass slides (Epredia^TM^ Superfrost^TM^Plus Adhesion Microscope Slides, J1820AMNZ, Thermo Fischer Scientific, USA). During cryosectioning, the glass slides were kept in the cryostat chamber during the sectioning to maintain RNA integrity. After sectioning, the slides were stored at −80°C until further processing.

For fibrotic tissue estimation, we stained for Fibronectin 1 (FN1). Glass slides were brought to RT and fixed in ice-cold acetone for 5 minutes after which they were air dried at RT for 30 minutes. A hydrophobic barrier was drawn using the ImmEdge™ Pen (H-4000, Vector Labs, CA, USA). Hereafter, sections were washed once using 1XPBS and blocked for 1 hour at RT using 1XPBS/0.05%Tween/5% horse serum. Subsequently, slides were incubated overnight with primary antibody for FN1 (Santa Cruz, Fibronectin (EP5), sc-8422, Mouse monoclonal, 1:400, diluted in 1XPBS/0.05%Tween/5%FBS) at 4°C. The next day, the slides were washed 3 times with 1XPBS and incubated for 60 minutes at RT with the Alexa 647 conjugated secondary antibody (Life Technologies, A21235, Goat-anti-mouse, Alexa 647, IgG, 1:500). Slides were then washed 3 times for 5 minutes with 1XPBS, mounted with DAPI (ProLongTM Gold antifade reagent with DAPI, P36935, Invitrogen, Thermo Fischer Scientific, Eugene, OR, USA) and covered with a cover slip. The slides were imaged on the ZEISS Axio Scan.Z1 slide Scanner (Carl Zeiss LTD, CAM, UK) with a 20x objective. DAPI was imaged in the 350nm channel and FN1 in the 674nm channel. The ZEN 2012 software (blue edition, Carl Zeiss LTD, CAM, UK) was used to process the images.

### Visium data pre-processing

Image analysis, base calling, and quality scoring of the sequencing data were performed with the Illumina data analysis pipeline RTA (v3.4.4) and the demultiplexed FASTQ data were generated by BClConvert (v3.10.5). We manually aligned the fluidic area and outlined the tissue in the histological image using Loupe Browser (v6.4.0). The alignment result with the FASTQ data were mapped using the “count” function in Space Ranger v2.0.1 with the GRCh38 human genome reference (refdata-gex-GRCh38-2020-A).

The data was analyzed with the Seurat package (v5.0.3) in R (v4.3.2). For each tissue section, we first manually identified the fat, folds and gaps based on the histological image as described above. Afterwards, we removed spots located in gaps, folds and spots with low counts that were not identified as fat (nSpatial_Count < 30 && label!= “fat”). Next, we filtered out all the mitochondrial and ribosomal genes. After filtering, all the datasets were normalized using log normalization with the NormalizeData function.

### Visium spot annotation using module scoring

To annotate the non-adipocyte spatial spots, we assigned marker genes to predefined modules (Connective tissue and Muscle Fibers) based on literature (Supplementary Table 2). We then calculated the module score for each defined module based on the average expression of these marker genes using the AddModuleScore function in Seurat. Based on the mixture distribution of the module scores for each module, we further divided the spatial spots for each sample into high- and low-level module score groups, with the threshold selected based on the majority votes from iterating the normalmixEM function from the mixtools R package (v2.0.0) 500 times.

First, spots within the high-level connective tissue module score group were assigned to the “Connective tissue” module. Then, the remaining spots were subdivided using the muscle fibers module score. Spots within the high-level muscle fibers module score group were assigned to the “Muscle Fibers” cluster, while the remaining spots were classified as “Not Assigned”.

To further subdivide spots within the “Muscle Fibers” module into different muscle fiber types, we compared the log-normalized expression levels for the myosin gene specific to the following muscle fiber types (Type I: *MYH7*; Type IIa: *MYH2*; Type IIx: *MYH1*). Each spot was further reassigned to a specific muscle fiber type based on the corresponding myosin gene with the highest expression level.

### Differential Gene Expression Analysis

Differential gene expression analysis was performed using the pseudo-bulk approach implemented in the muscat R package (v1.13.1). We conducted comparative analysis between different conditions (HC vs. BMD, HC vs. DMD, BMD vs. DMD) at both the sample-level and the module-level.

At the sample-level, pseudo-bulk profiles were generated for each individual sample by aggregating counts across all spots. For the module-level comparison, pseudo-bulk profiles were created by summing counts for spots within each annotated cluster per sample for each condition.

Pseudo-bulk profiles with fewer than 10 cells were excluded from the analysis. Additionally, lowly expressed genes were filtered out using filterByExpr function with the default setting in the edgeR package (v3.42.4). Next, differentially expressed genes (DEGs) across conditions were identified using the edgeR method in muscat, with the selection cutoff |log_2_(fold-change)| > 0.5 and adjusted *P*-value < 0.05 by Benjamini-Hochberg procedure.

### Deconvolution

We used the CARD R package (v1.1) to deconvolute the Visium data and infer the cell type composition for each spot. CARD is based on a non-negative matrix factorization model, utilizing the cell type markers from a single-nuclei RNA sequencing (snRNAseq) reference dataset for deconvoluting spatial transcriptomics data. Besides, CARD also leverages spatial information to enable accurate deconvolution in spatial transcriptomics. For the reference data, we used snRNAseq from Suárez-Calvet et al., which includes human skeletal muscle from healthy and DMD individuals (Suárez-Calvet et al., 2023). First, we removed all the mitochondrial and ribosomal genes from the snRNAseq data. Next, we excluded the ‘B/T cells’ cell type from the reference data because there were too few cells (*N* = 188) to obtain reliable estimates for this cell population. Then we identified marker genes of the remaining nine cell types (Fast fibers, Slow fibers, Regenerative fibers, Adipocytes, FAPs, Endothelial cells, Satellite cells, Smooth muscle cells and Monocytic cells) using Seurat::FindAllMarkers (min.pct = 0.1, logfc.threshold = 2.5, only.pos = TRUE), and selected the top 50 marker genes per cell type under the threshold of adjusted *P*-value < 0.05 based on Bonferroni correction as the input for CARD deconvolution.

### Cell-cell communication analysis

LIANA+ (lianapy v0.1.8; Dimitrov et al., 2024) was used to compute cell-cell communication scores between cell types surrounding adipose tissue. To study distance-dependent communication events, we established three adipose-adjacent layers manually. We specified a combination of NMD status, sample of origin and layer as the *sample_context* for the spots used as input to *liana.method.rank_aggregate.by_sample* function. The cell type assignments after deconvolution were used as the group option. LIANA’s Consensus database was used as a ligand-receptor resource. We applied two filtering steps to identify relevant ligand-receptor interactions: first, we excluded pairs that fell below an expression threshold of 0.6 (i.e., retaining only interactions where both ligand and receptor were expressed in ≥ 60% of spots for any given cell type pair); second, we compared the expression levels between the adipose-adjacent layers and the remaining muscle tissue. Only ligand-receptor pairs showing at least a two-fold higher mean expression in the adipose-surrounding layers were retained for further analysis.

To capture cell-cell communication trends along the layers we used Tensor-Cell2Cell v0.6.8 (Armingol et al., 2022). Briefly, we reduced the dimensionality of the computed communication scores using tensor factorization. The input tensor contained the cell-cell communication scores between pairs of cell types in each sample context (i.e. in which layer and sample each spot is located) and for each ligand and receptor pair. In total, 21 contexts were defined across 7 cell types and 649 ligand-receptor pairs, which resulted in a tensor with dimensions 21×649×7×7. After factorization, we obtained 15 factors of cell-cell communication trends across each of the original dimensions in 4 different matrices. For the factorization, the *cell2cell.analysis.run_tensor_cell2cell_pipeline* function was used with robust optimization, random initialization, and NumPy’s Singular Value Decomposition (SVD) solver.

### Spatial fat analysis

We applied SPATA2 (v3.0.0) to identify genes whose expression patterns are influenced by fat tissue. For samples containing fat areas, we imported the space ranger output to SPATA2 object by using the command: SPATA2::initiateSpataObjectVisium(). Next, we annotated the fat areas based on the HE stained images by manually outlining their boundaries using SPATA2::createImageAnnotations(). We then applied the function SPATA2::spatialAnnotationScreening() to screen for genes with non-random patterns along the distance from the annotated fat areas to all tissue spots. Only spatially variable genes with non-zero counts were included in the analysis based on the function SPATA2:: removeGenesZeroCounts followed with SPATA2:: getSparkxGenes(…, threshold_pval = 0.05). Pattern genes associated with the fat trajectory were selected based on adjusted *P*-value < 0.05 based on Benjamini-Hochberg correction and Root Mean Squared Error (RMSE) < 0.25, fitting with the descending or ascending models predefined by SPATA2.

### Statistics

To assess significant differences in module annotation percentages and cell type percentages from deconvolution, linear models testing the percentages of modules and cell types across samples in the cohorts (HC, BMD and DMD) with the HC samples as a reference were tested without interaction. Moreover, the same models were tested with BMD as a reference to test the differences between BMD and DMD. The outcomes of these linear models are summarized in Supplementary Table 4 and 7.

## Data and code availability

Spatial transcriptomics count data will be available on a public server upon publication. All original code has been deposited on GitHub https://github.com/Qirongmao97/NMDhuman_spatial) and is publicly available as of date of publication. The used snRNAseq data is available upon reasonable request through the corresponding author of that dataset (Suárez-Calvet et al., 2023).

## Author contribution

L.H. performed the wet lab experiments, parts of the analysis and drafted the first version of the manuscript. Q.M. performed parts of the analysis and wrote parts of the manuscript. S.F. and K.F. provided biopsies of a healthy control, four BMD and three DMD samples and allowed for collaborative execution of the wet lab experiments by L.H. in their lab. C.N.R. performed the CCC analysis and wrote the methods on this part. Under supervision of L.H., J.v.d.W. performed the immunofluorescent staining on the skeletal muscle. J.K. contributed to the implementation of the SPATA2 pipeline for the skeletal muscle tissue. E.N. and H.K. provided biopsies of healthy controls and one DMD sample from Leiden University Medical Center and The Hague. J.M.D. and R.G.N. provided the snRNAseq dataset. P.S., A.M., M.v.P. and A.A.R. supervised the work, led by P.S. and A.M.. All authors provided feedback and comments on the manuscript draft.

## Acknowledgements

Healthy control biopsies from the Netherlands were supported the Netherlands Organization for Scientific Research (NWO, under research program VIDI, Grant # 917.164.90)

