## Supplementary for "Unraveling the spatial landscape of Dystrophinopathies: a transcriptomic approach to Becker and Duchenne muscular dystrophies"

 **Affiliations:**
^1^Department of Human Genetics, Leiden University Medical Center, The Netherlands
^2^Center for Gene Therapy, The Abigail Wexner Research Institute at Nationwide Children's Hospital, Columbus, OH 43205, USA
^3^Microenvironment and Immunology Research Laboratory, Medical Center, Faculty of Medicine, Freiburg University, Freiburg, Germany
^4^Department of Neurosurgery, Medical Center, Faculty of Medicine, Erlangen University, Erlangen, Germany
^5^John Walton Muscular Dystrophy Research Centre, Newcastle University Translational and Clinical Research Institute, Newcastle Upon Tyne NHS Trust, Newcastle Upon Tyne, United Kingdom
^6^Laboratori de Malalties Neuromusculars, Institu de recerca Hospital Sant Pau, Barcelona, Spain
^7^Centro de Investigación Biomédica en red en enfermedades raras (CIBERER)
^8^Department of Radiology, Leiden University Medical Center, Leiden, The Netherlands
^9^Duchenne Center Netherlands

^10^Department of Neurology, Leiden University Medical Center, Leiden, The Netherlands

^11^Department of Pediatrics, The Ohio State University, Columbus, OH 43210, USA
^12^Department of Neurology, The Ohio State University, Columbus, OH 43210, USA
^13^Delft Bioinformatics Lab, Delft University of Technology, The Netherlands

*Authors contributed equally

**ORCIDs:**
Laura GM Heezen	0000-0002-0789-0347
Qirong Mao	0000-0001-7938-6660
Claudio Novella Rausell	0000-0002-7383-6090
Annemieke Aartsma-Rus	0000-0003-1565-654X
Pietro Spitali	0000-0003-2783-688X
Ahmed Mahfouz	0000-0001-8601-2149
Maaike van Putten	0000-0002-0683-8897
Erik H Niks	0000-0001-5892-5143
Hermien E Kan	0000-0002-5772-7177
Stefan Nicolau	0000-0001-5631-0559
Kevin Flanigan	0000-0001-6440-3376
Jordi Diaz Manera	0000-0003-2941-7988

**Abstract**

Dystrophinopathies are caused by pathogenic variants in the *DMD* gene resulting in partial (Becker) or complete loss (Duchenne) of dystrophin. Becker (BMD) and Duchenne muscular dystrophy (DMD), are characterized by progressive muscle wasting, fatty replacement, fibrosis, and loss of function. To study histopathological changes, we used spatial transcriptomics to profile skeletal muscle biopsies of BMD, DMD patients and healthy controls (*N* = 4 per group). We estimated the proportion of cell types and their spatial localization across samples applying a deconvolution strategy using single-nuclei RNA-sequencing data. We identified genes enriched in fat patches and cell types such as fibroadipogenic progenitor cells (FAPs) in areas of active pathology. Using expression data of ligand receptor pairs, we highlight cell-cell communications leading to fibrotic and adipogenic lesions. Finally, analysis of gene expression gradients in areas of adjacent muscle and fat, allowed the identification of genes associated with muscle areas committed to become fat.

*Keywords*: spatial transcriptomics, dystrophinopathy, fibroadipogenic progenitors, histopathology, FAPs, cell-cell communication, single nuclei RNA sequencing

**Supplemental information**

**Table 1:	Technical details on Visium Spatial Gene Expression slide processing
Table 2:	Genes used for module scored annotation**

**Figure 1:	all HE images of samples included in the study
Figure 2:	FN1 stained HC samples
Figure 3:	FN1 stained BMD samples
Figure 4:	FN1 stained DMD samples**

**Table 3:	Module annotation numbers per sample for all modules
Table 4:	Linear model outcomes module annotation**

**Table 5:	DGE across diseases and modules
Figure 5:	DMD expression spatially mapped**

**Table 6:	Deconvolution numbers per sample for all cell types
Table 7:	Linear model outcomes cell type**

**Table 8:	CCC results 0.6 resolution
Figure 6:	CCC BMD layers & cell types
Figure 7:	CCC DMD layers & cell types
Figure 8-9:	CCC top L-R pairs across samples**
**Figure 10:**	**Markers subset of interesting FAPs based on CCC analysis**

**Figure 11:	Adipogenic marker genes across samples
Table 9:	Genes with informative spatial patterning detected with SPATA2
Figure 12:	SPATA2 plot genes across samples**

**Table 1:	Technical details on Visium Spatial Gene Expression slide processing**

| **Cohort** | **Sample** | **Slide number** | **Capture area position** | **% tissue coverage** | **cDNA cycles amplification** | **SI PCR cycle number** |
| --- | --- | --- | --- | --- | --- | --- |
| HC | 1 | V19N11-029 | A | 40 | 16 | 18 |
|  | 2 | V12Y31-101 | C | 25 | 19 | 18 |
|  | 3 | V11D13-059 | D | 40 | 23 | 14 |
|  | 4 | V11D13-059 | B | 67 | 20 | 13 |
| BMD | 1 | V12Y31-125 | B | 35 | 17 | 15 |
|  | 2 | V12Y31-125 | D | 19 | 17 | 16 |
|  | 3 | V12Y31-125 | A | 23 | 16 | 16 |
|  | 4 | V12Y31-101 | B | 36 | 17 | 16 |
| DMD | 1 | V19N11-029 | B | 18 | 16 | 18 |
|  | 2 | V12Y31-125 | C | 33 | 16 | 16 |
|  | 3 | V12Y31-101 | D | 28 | 19 | 18 |
|  | 4 | V12Y31-101 | A | 43 | 16 | 15 |

*Supplementary Table* *1*. Technical details on Visium slide number, position of capture area and amount of PCR cycles used in Visium Spatial Gene Expression experiments. SI; sample index.

**Table 2:	Genes used for module scored annotation**

| **Connective tissue** | **Muscle Fibers** | **Type I** | **Type IIa** | **Type IIx** |
| --- | --- | --- | --- | --- |
| *COL1A1* | *CKM* | *MYH7* | *MYH2* | *MYH1* |
| *COL1A2* | *TNNT3* |  |  |  |
| *THBS4* | *TPM1* |  |  |  |
| *COL3A1* | *TPM2* |  |  |  |
| *FN1* | *TNNT1* |  |  |  |
| *COL6A6* | *MYH1* |  |  |  |
| *DCN* | *MYH2* |  |  |  |
| *GSN* | *MYH7* |  |  |  |
| *BGN* | *ATP2A1* |  |  |  |
|  | *TNNI2* |  |  |  |
|  | *ENO3* |  |  |  |
|  | *PFKM* |  |  |  |
|  | *PKM* |  |  |  |
|  | *TNNI1* |  |  |  |
|  | *MYH7B* |  |  |  |

*Supplementary Table* *2*. Genes used for module scored annotation for sample.

**Supplementary Figure 1:	all HE images of samples included in the study**

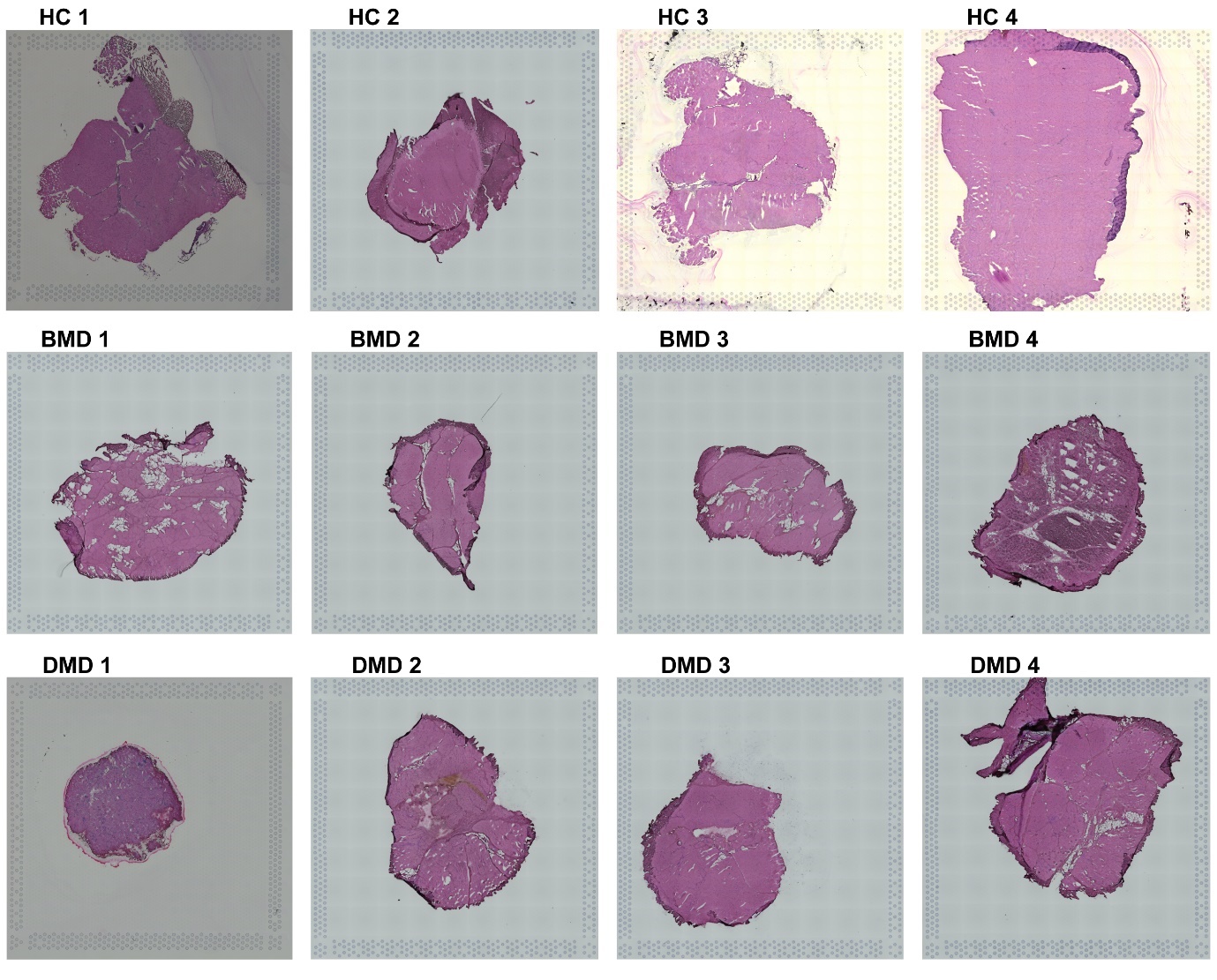

Supplementary Figure 1. **All HE stained samples included in the study.** HC; healthy control, BMD; Becker muscular dystrophy, DMD; Duchenne muscular dystrophy.

**Supplementary Figure 2:	FN1 stained HC samples**

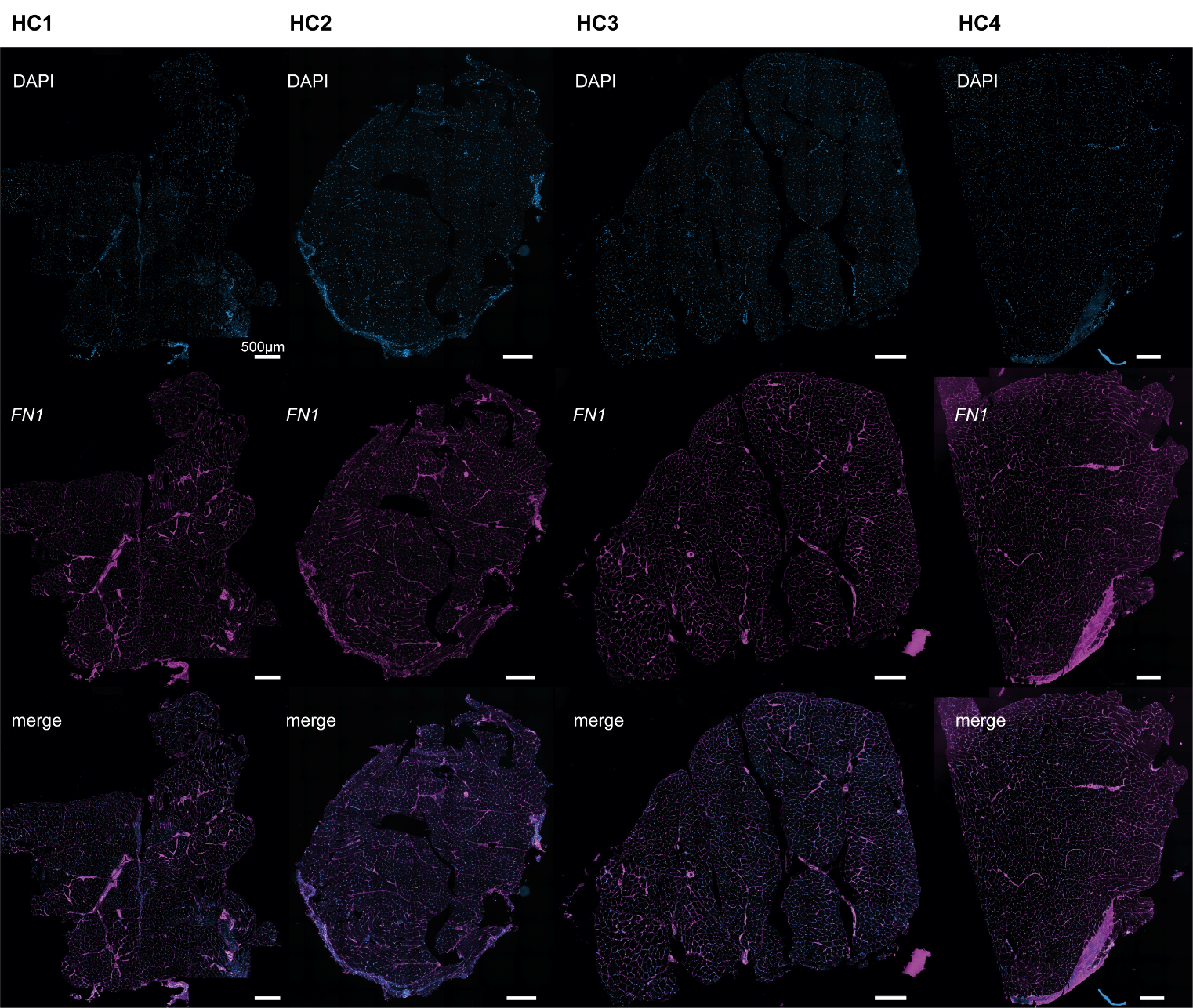

Supplementary Figure 2. **All healthy controls (HC) stained for FN1 as a marker of fibrosis and connective tissue co-stained with DAPI highlighting the nuclei.**

**Supplementary Figure 3:	FN1 stained BMD samples**

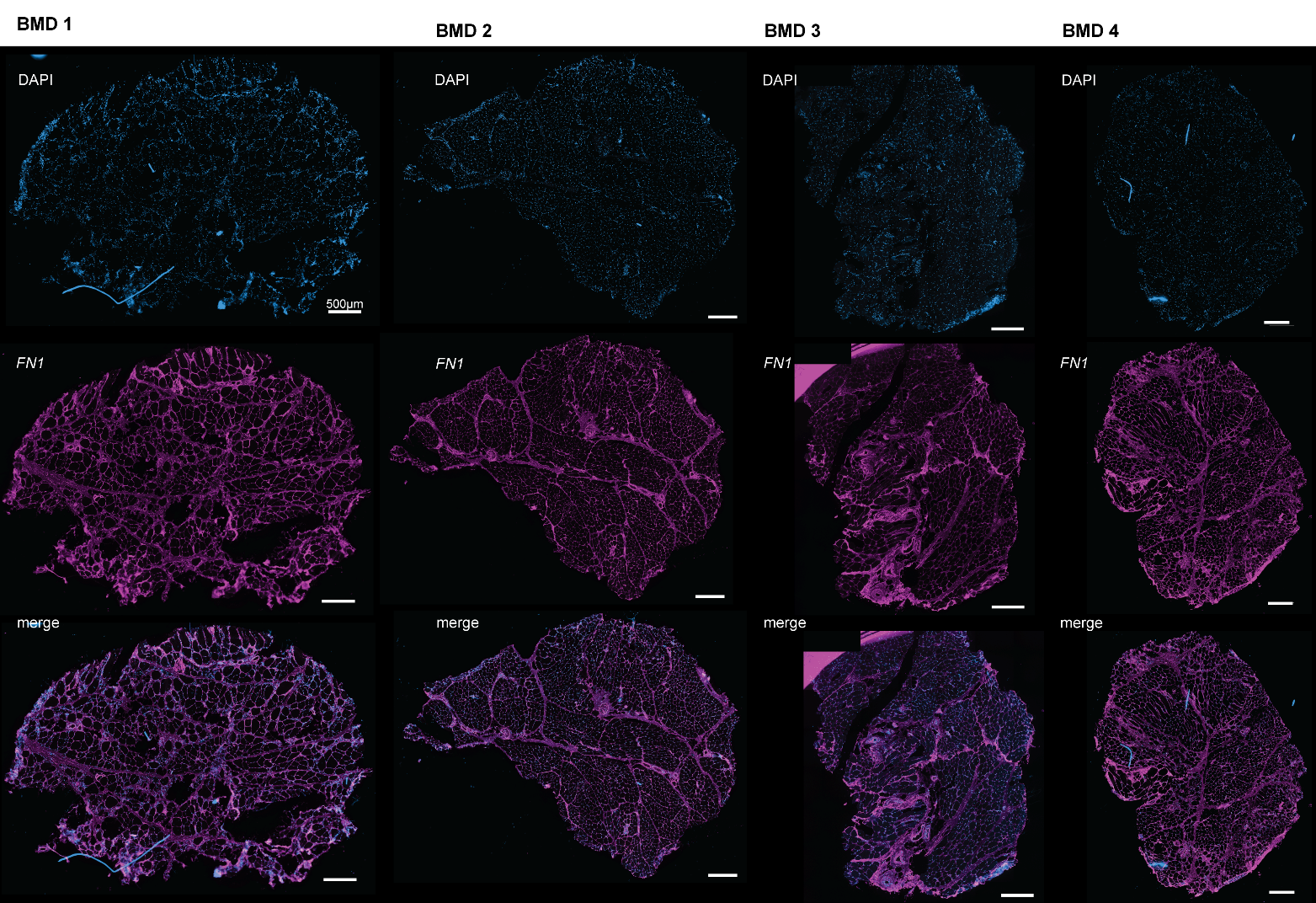

Supplementary Figure 3. **All BMD samples stained for FN1 as a marker of fibrosis and connective tissue co-stained with DAPI highlighting the nuclei.**

**Supplementary Figure 4:	FN1 stained DMD samples**

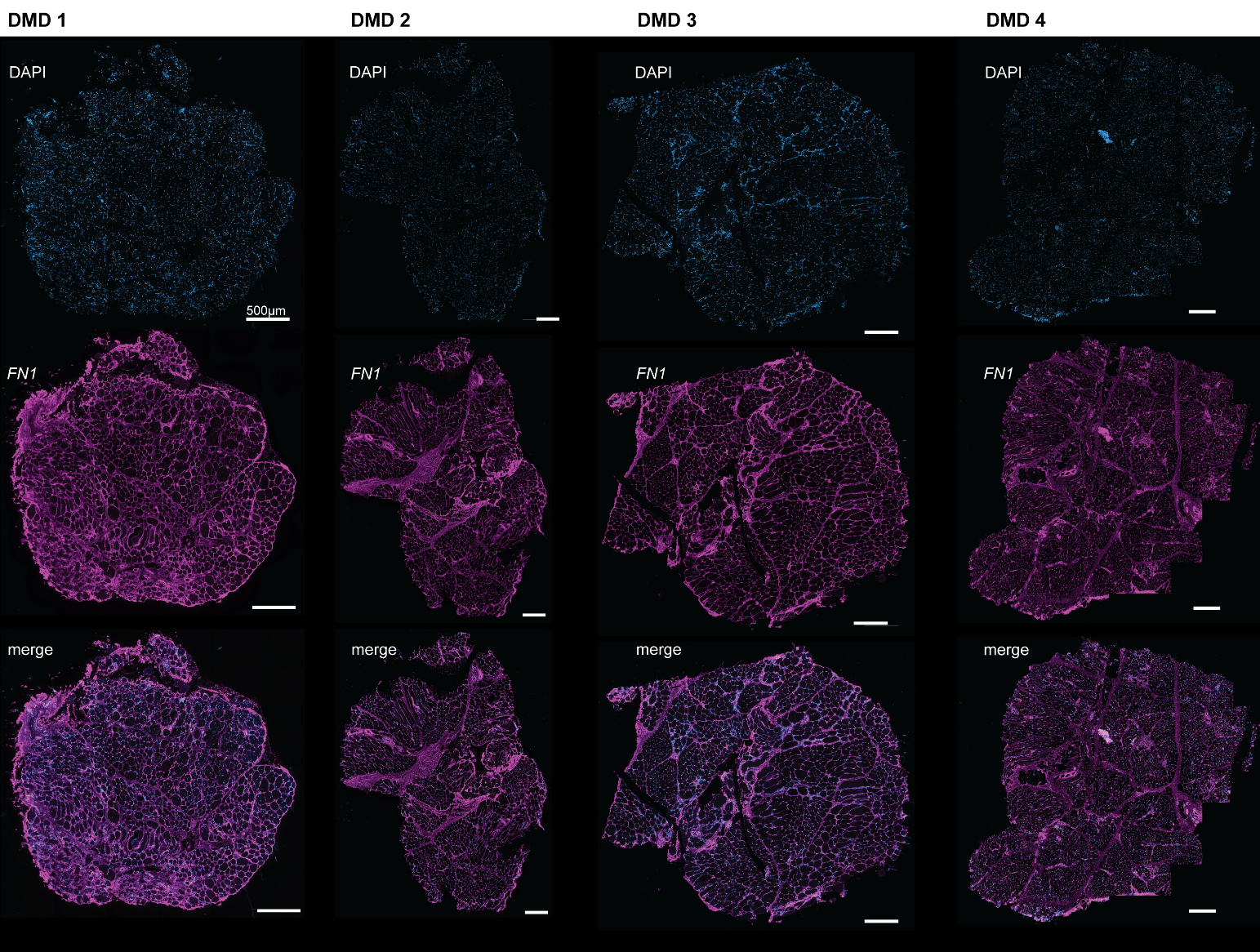

Supplementary Figure 4. **All DMD samples stained for FN1 as a marker of fibrosis and connective tissue co-stained with DAPI highlighting the nuclei.**

**Supplementary Figure 5:	DMD expression spatially mapped**

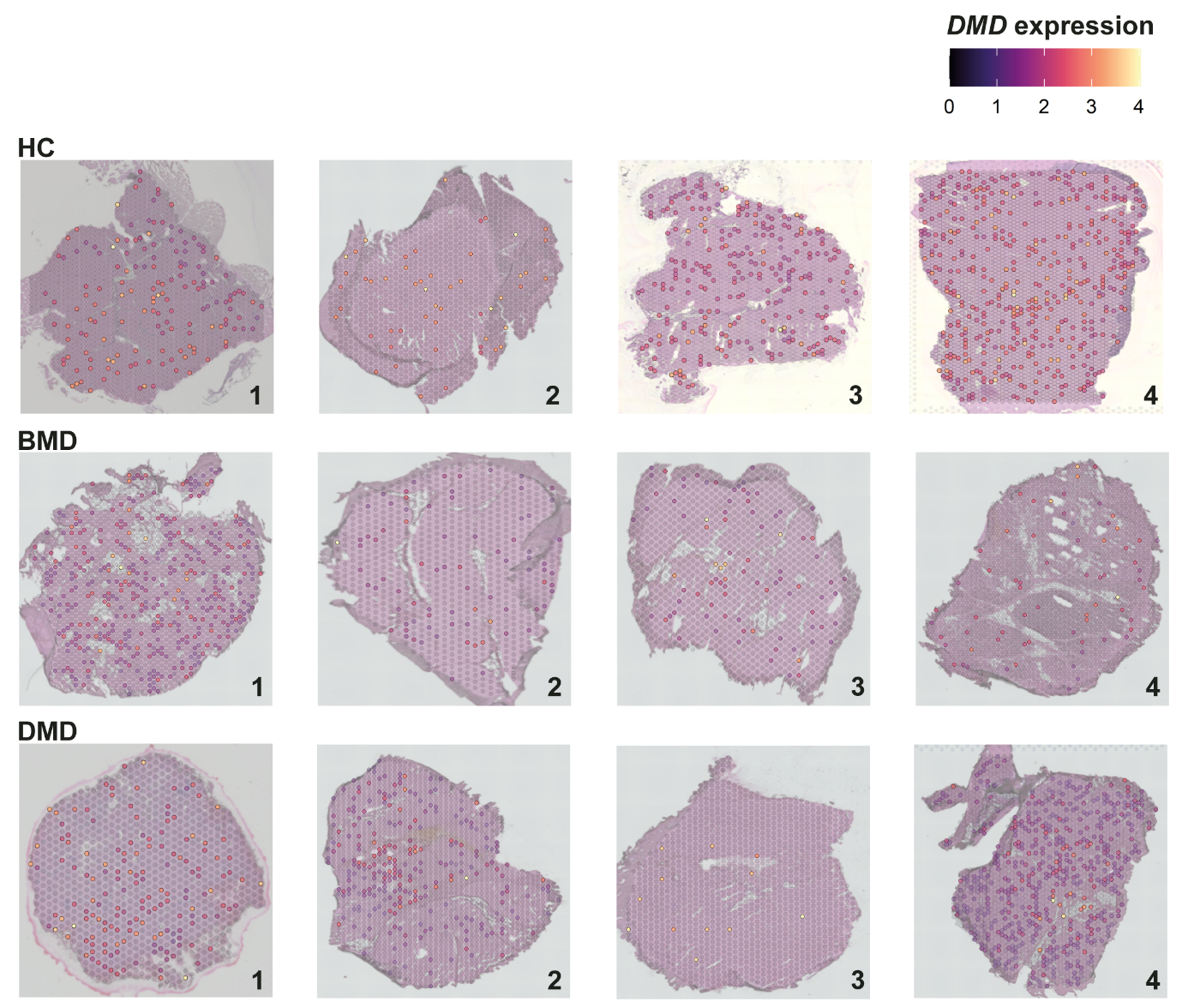

Supplementary Figure 5. ***DMD* expression spatially plotted across all samples included in the study.**

**Supplementary Figure 6:	CCC BMD layers & cell types**

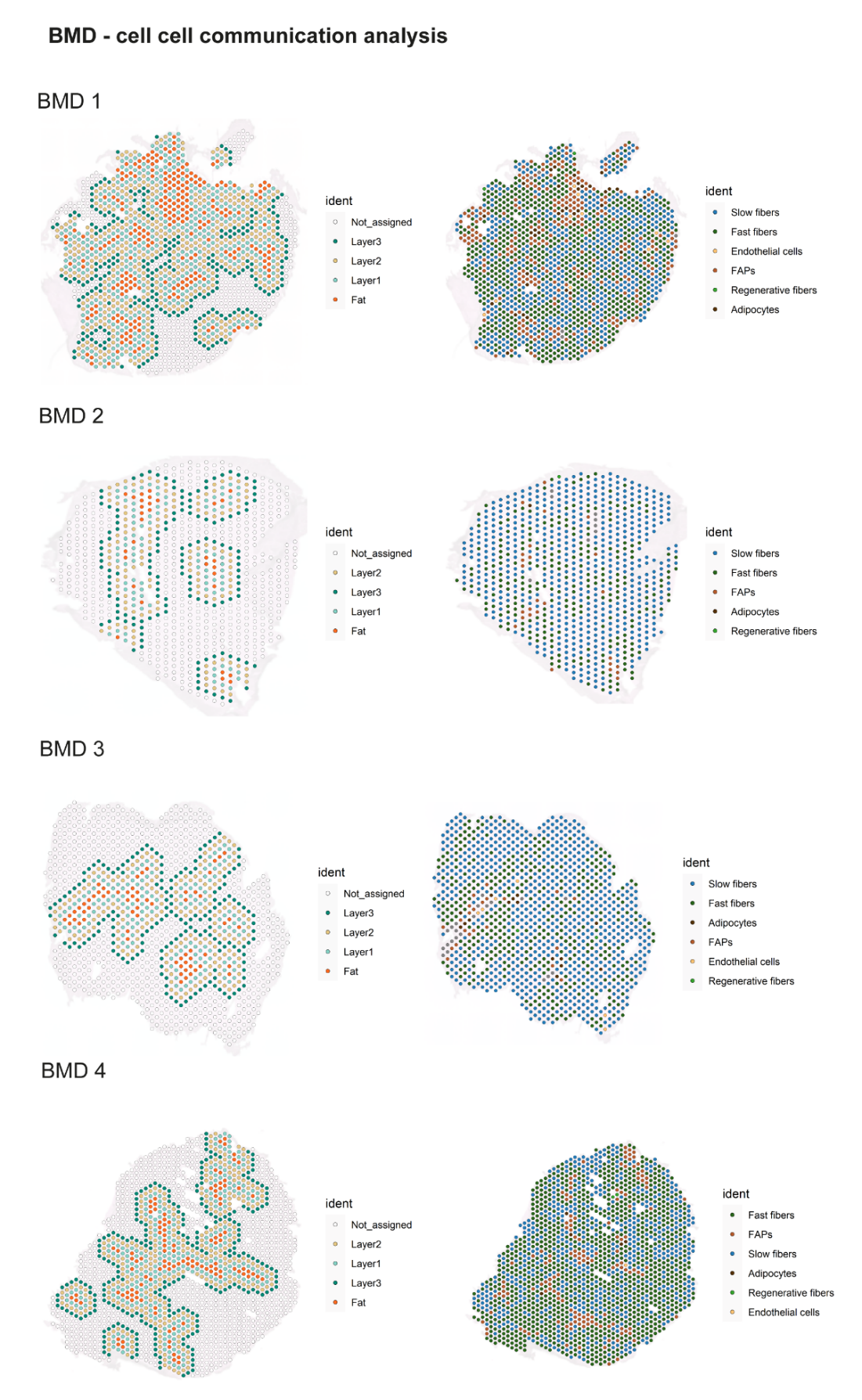

Supplementary Figure 6. **BMD samples included in CCC analysis, fat and surrounding layers annotated included into the analysis as well as the assigned cell type.**

**Supplementary Figure 7:	CCC DMD layers & cell types**

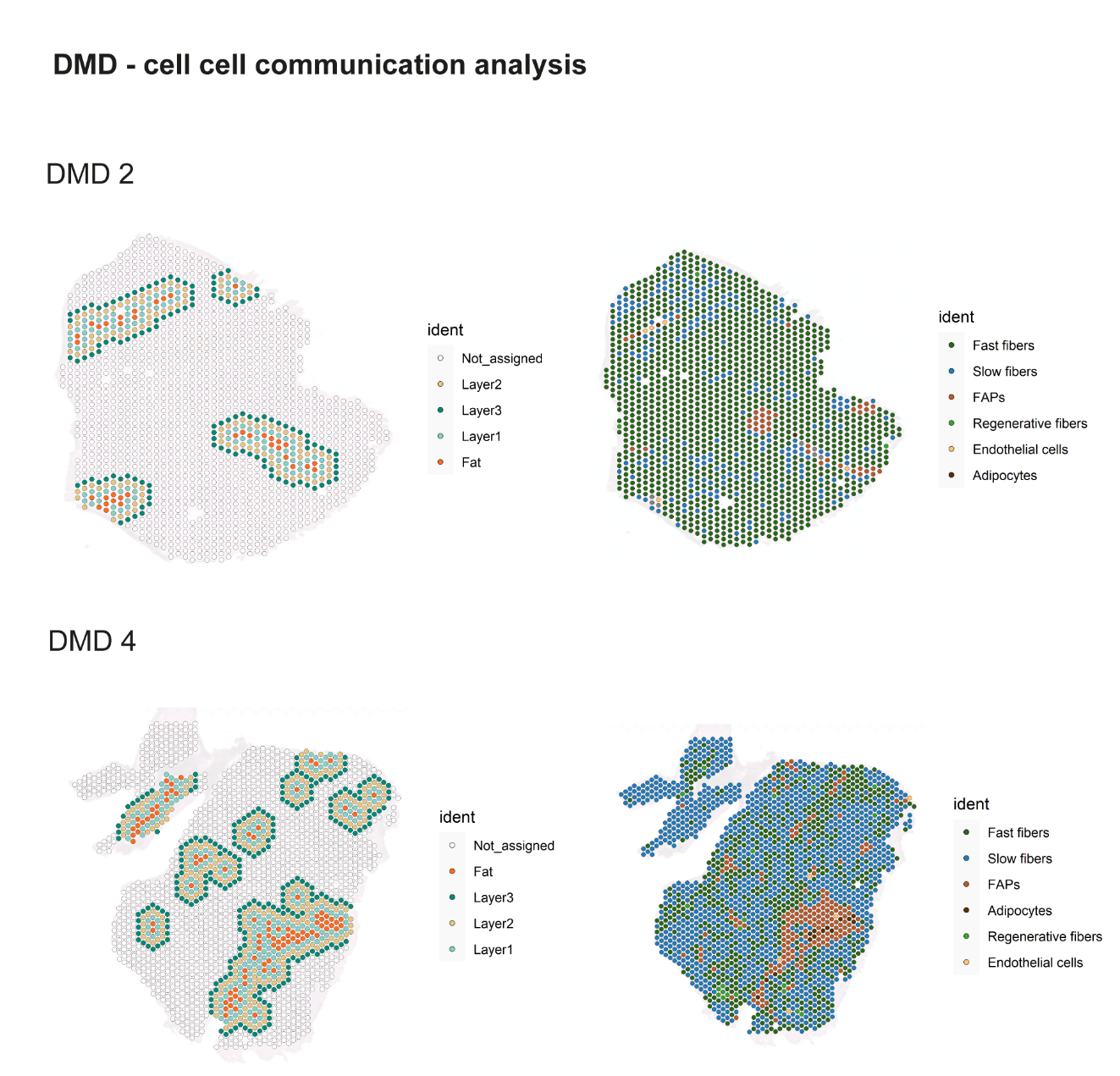

Supplementary Figure 7. **DMD samples included in CCC analysis, fat and surrounding layers annotated included into the analysis and the assigned cell type.**

**Supplementary Figure 8-9:	CCC top L-R pairs across samples**

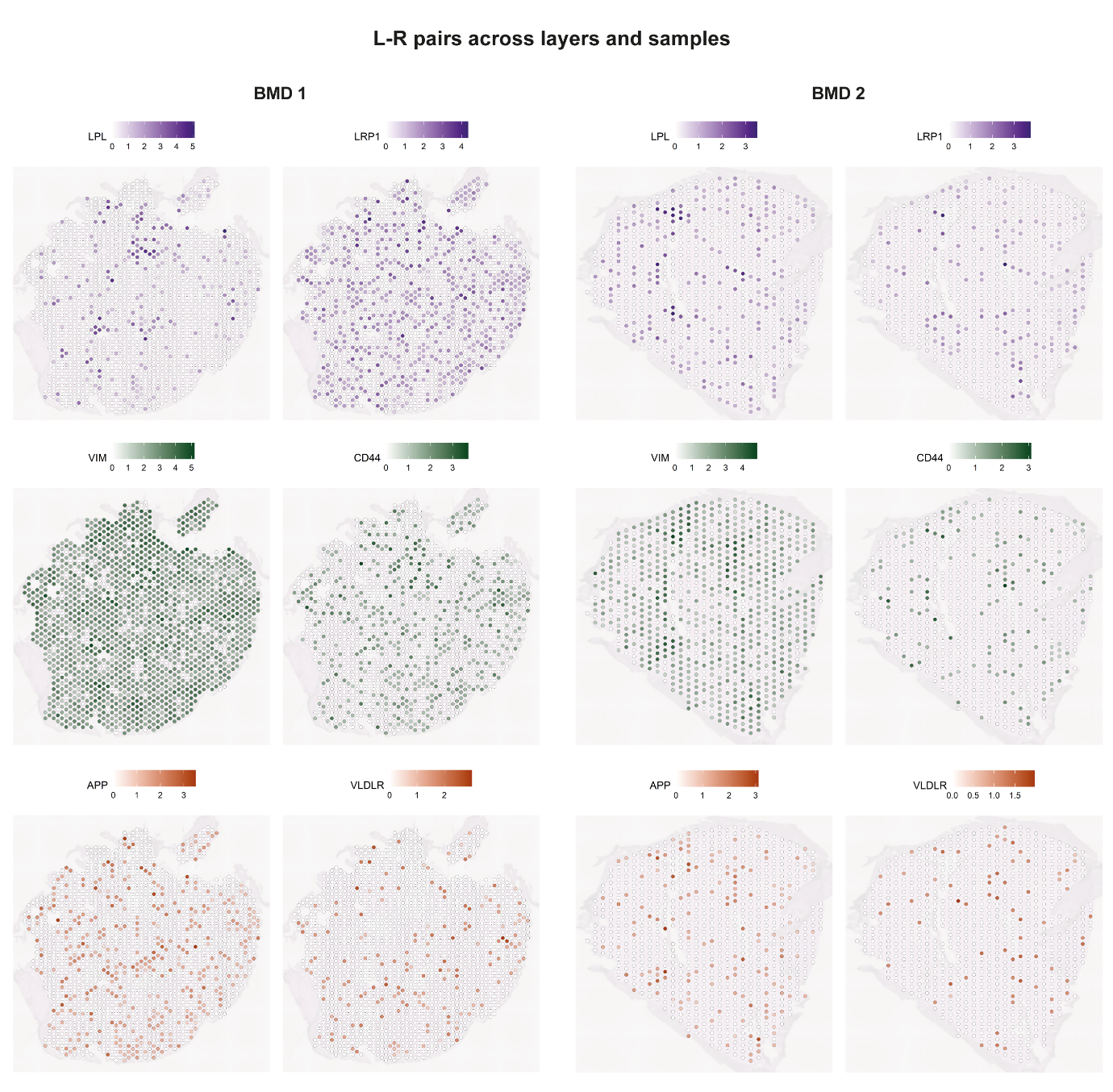

Supplementary Figure 8. **L-R pairs detected across layers and plotted across samples (BMD 1 and BMD 2).**

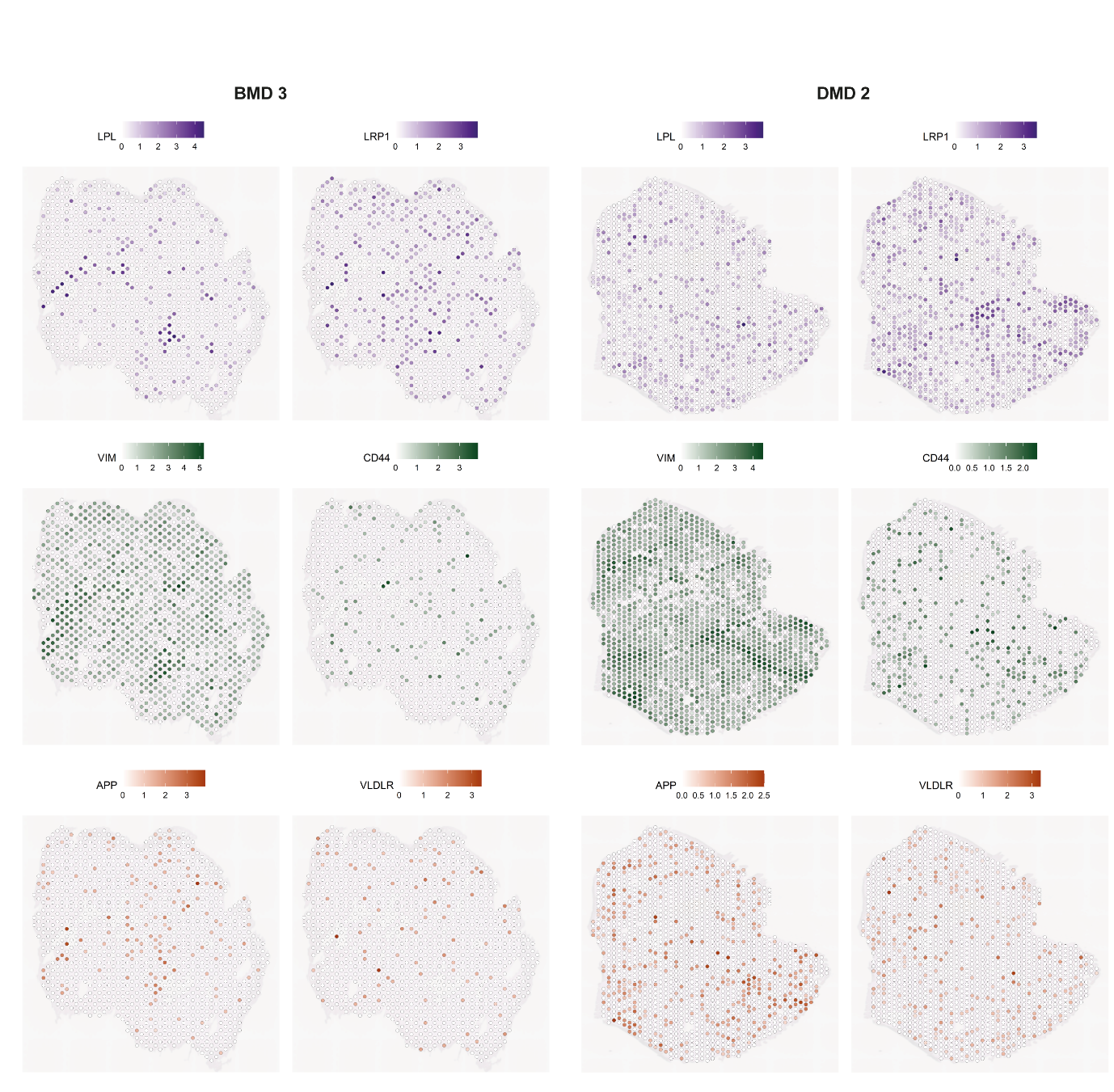

Supplementary Figure 9. **L-R pairs detected across layers and plotted across samples (BMD 3 and DMD 2).**

**Supplementary Figure 10:	Markers subset of interesting FAPs based on CCC analysis**

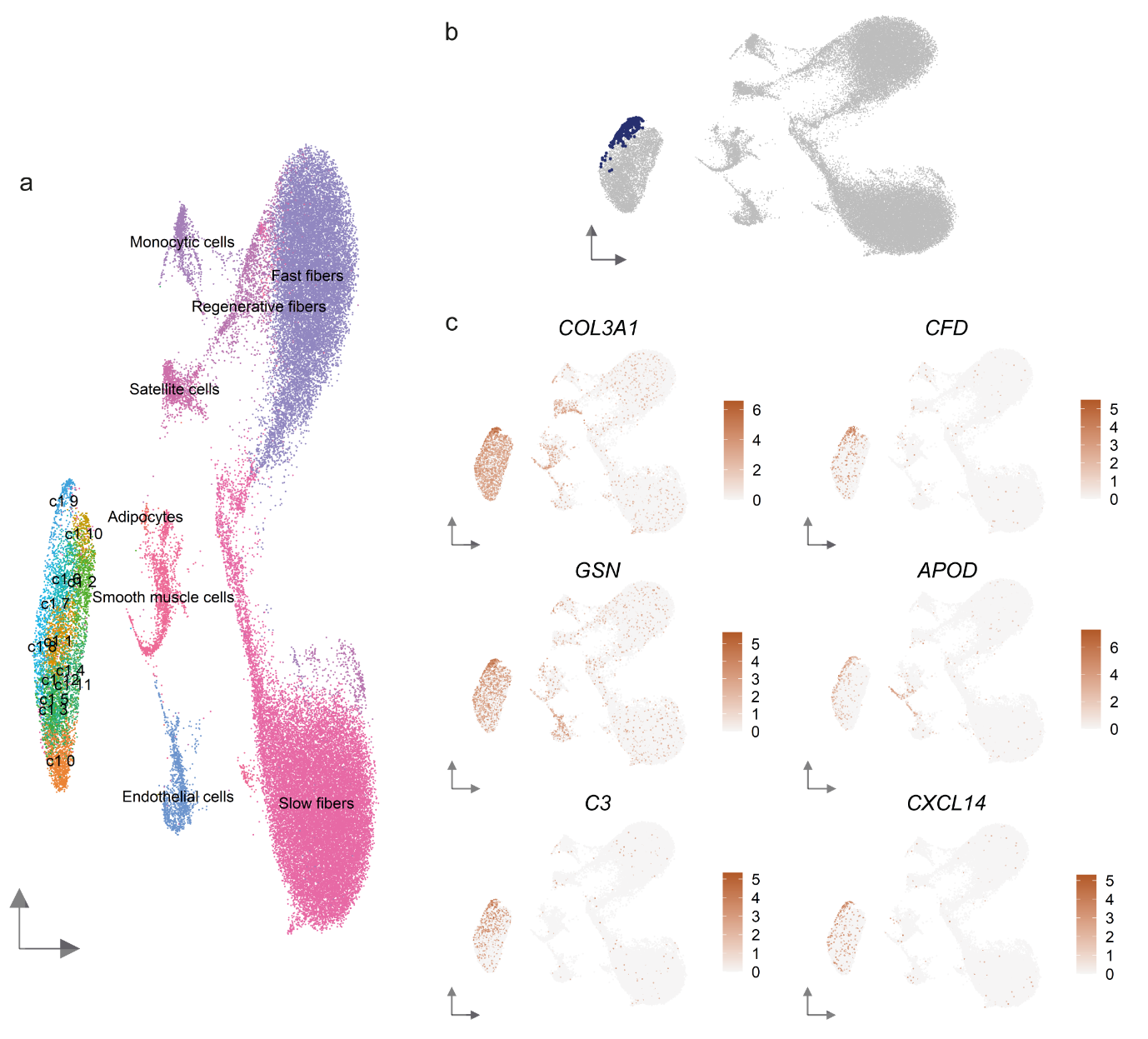

Supplementary Figure 10. **Reclustering FAPs from single nuclei dataset to detect markers of specific FAP cluster thought to be representative of the FAPs in our CCC analysis.** (a) Reclustering of the FAP cohort from the single cell dataset divides the FAPs into multiple clusters. (b) Cluster c19 is the main cluster expressing L-R markers from our CCC analysis. (c) A few top expressing markers for this specific cluster are *COL3A1, CFD, GSN, APOD, C3* and *CXCL14*.

**Supplementary Figure 11:	Adipogenic marker genes across samples**

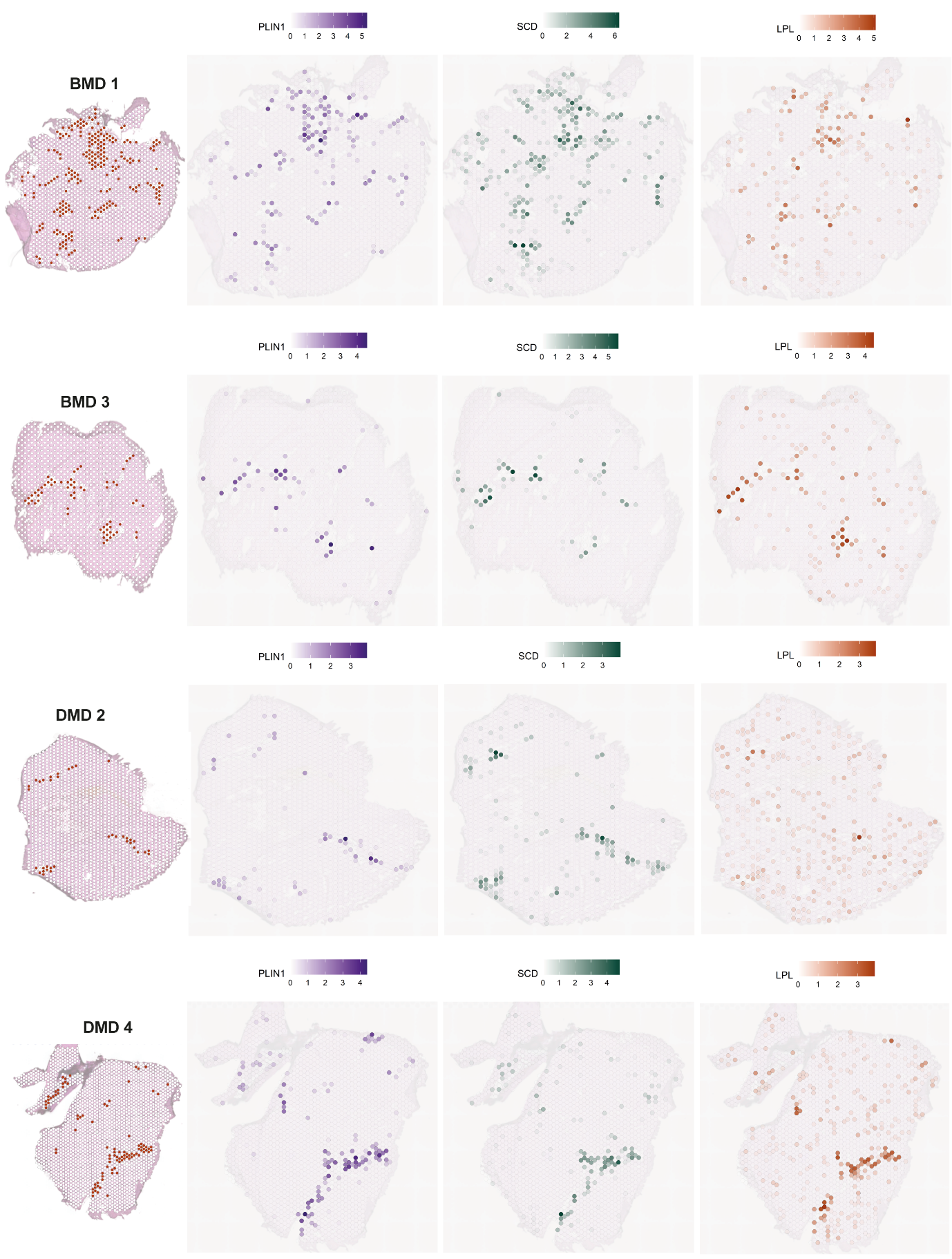

Supplementary Figure 11. **Spatial mapping of a few adipogenic marker genes (*PLIN1*, *SCD*, *LPL*) across samples included in the spatial fat analysis.**

**Supplementary Figure 12:	SPATA2 plot genes across samples**

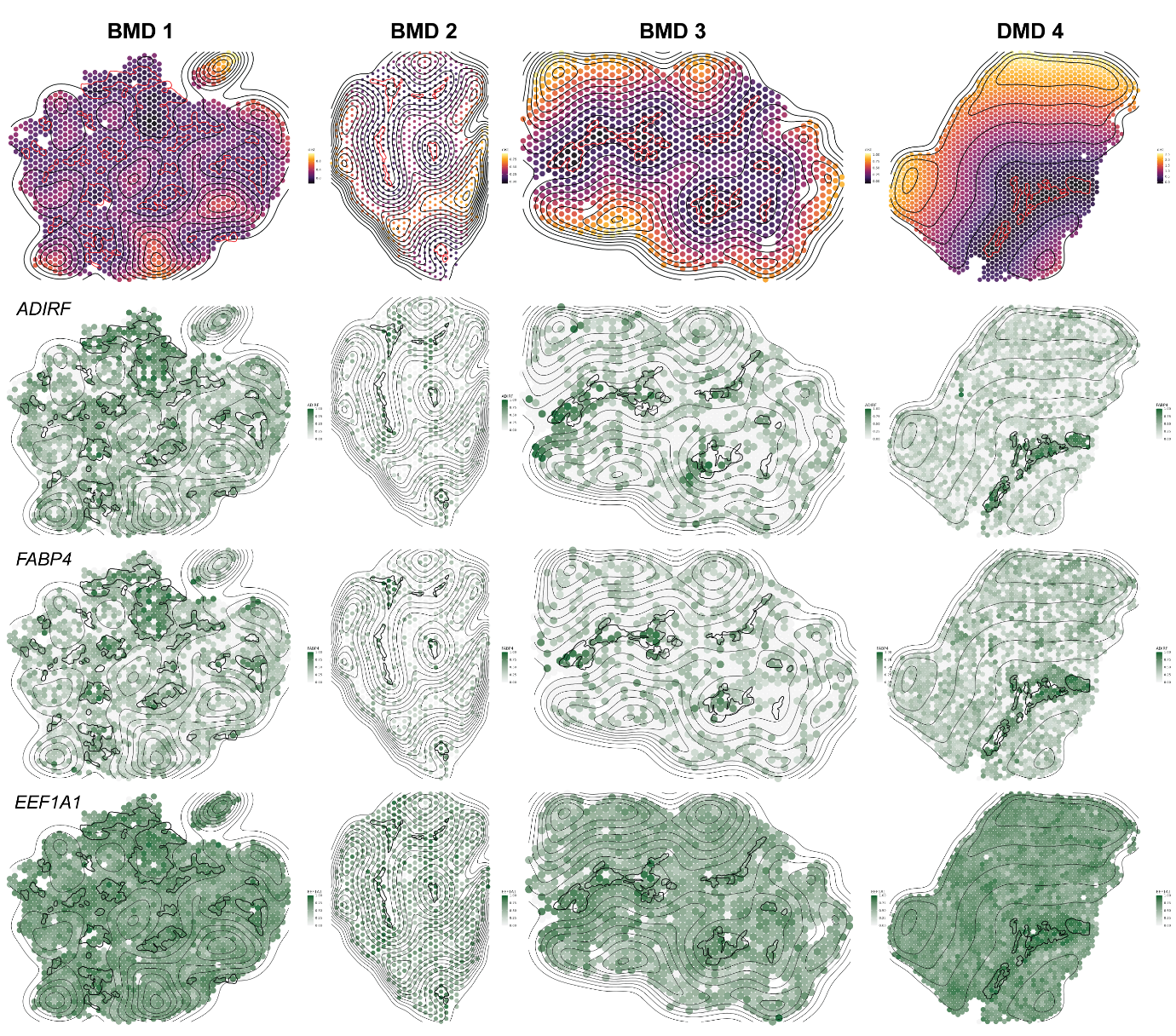

Supplementary Figure 12. **A selection of three SPATA genes (*ADIRF, FABP4, EEF1A1*) plotted spatially across samples.**
